# PATHOS: Predicting Variant Pathogenicity by Combining Protein Language Models and Biological Features

**DOI:** 10.64898/2025.12.22.25342839

**Authors:** Ragousandirane Radjasandirane, Gabriel Cretin, Julien Diharce, Alexandre G. de Brevern, Jean-Christophe Gelly

## Abstract

Predicting the pathogenic impact of missense variants is essential for understanding and diagnosing genetic diseases. These approaches have undergone significant evolution, with the latest methodologies based on deep learning approaches. Nonetheless, only a limited number use the potential of Protein Language Models (PLMs), which have demonstrated strong performance across various protein-related tasks.

A new predictor, called PATHOS, was developed; it combines embeddings from an optimal set of two PLMs, namely ESM C 600M and Ankh 2 Large. Their embeddings were combined with additional crucial biological features such as phylogenetic probabilities, allele frequency, and protein annotations; they were aggregated using a fully connected layer architecture.

Compared to 65 other predictors on clinical data, PATHOS outperforms state-of-the-art performance. It achieves a Matthews Correlation Coefficient (MCC) of 0.591 on a manually and carefully curated clinical dataset and 0.826 on a ClinVar dataset, surpassing other leading tools. Furthermore, case studies on the progesterone receptor and the KCNQ1 ion channel illustrate that PATHOS can identify functionally critical regions and known pathogenic mutations missed by other leading predictors like AlphaMissense.

To ensure broad accessibility and facilitate use by non-specialists, a user-friendly web server containing a database of 140 millions precomputed predictions from human protein from Swiss-Prot was provided. The web server is available at: https://dsimb.inserm.fr/PATHOS/

## Introduction

In a cell, DNA replication is not entirely error-free even with sophisticated correction mechanisms (Kunkel & Bebenek, 2000). These infrequent errors can result in genetic diseases (Cooper et al., 2010; Subramanian, 2012). If the mutation is located in germinal cells, this mutation will likely be inherited by the next generation, possibly resulting in an inheritable genetic disease. The sequencing of numerous human genomes showed that our DNA is frequently subject to nucleotide changes (The 1000 Genomes Project Consortium et al., 2015). These changes range from Single Nucleotide Variation (SNV), to deletions, insertions, combinations of both going up to chromosome rearrangements. Most nucleotide changes are harmless and only a small part of them have been linked to known diseases (Landrum et al., 2014; Stenson et al., 2003), i.e. Benign and Pathogenic mutations. Distinguishing pathogenic mutations from benign ones is essential to rapidly and effectively identifying the genetic disease that affects the patient, thereby guiding its diagnosis and potential treatments (Richards et al., 2015). Different techniques may be employed by clinicians or medical geneticists to this end, but they usually require time and are often expensive. (van Nimwegen et al., 2015).

Over the past two decades, researchers have developed a tremendous number of methods, called Variant Effect Predictors (or VEPs), to predict the impact of mutations (Livesey & Marsh, 2020, 2025; Radjasandirane et al., 2025; Riccio et al., 2024). The pioneer VEPs were SIFT (Ng & Henikoff, 2003) and PolyPhen (Adzhubei et al., 2010, 2013); they were based on simple principles such as residue conservation through evolution. These VEPs were widely used in the past and continue nowadays. At the same time, numerous new VEPs have been developed with more complex algorithms that clearly outperform SIFT and PolyPhen. Machine Learning, and especially Deep Learning based VEPs (such as EVE (Frazer et al., 2021), AlphaMissense (Cheng et al., 2023) or CPT-1 (Jagota et al., 2023)), have significantly improved performances. Nonetheless, a room for improvement still exists since current best VEPs are far from the required performances to allow a safe and reliable use of VEP tools in a clinical context. Currently, the American College of Medical Genetics and Genomics (ACMG) and the Association for Molecular Pathology (AMP) ranked VEPs as the weakest criterion to clinically classify a new variant (Richards et al., 2015).

The use of general-purpose PLMs is a promising way to increase predictions accuracy of the pathogenicity of mutations, as they already prove to capture a deep understanding of relationships between residues within a protein sequence (J.-Y. Chen et al., 2025; Elnaggar et al., 2022). This understanding also encompasses structural and functional information relating to the protein sequence. This information can be useful for predicting pathogenicity. Here, the performance of nine PLMs were analyzed in the pathogenicity prediction task. Our research includes PLMs from Rost’s Lab namely Ankh models (Elnaggar et al., 2023) (Ankh Base, Ankh Large, Ankh Large 2) and T5-based models (ProtT5 (Elnaggar et al., 2022), and ProstT5 (Heinzinger et al., 2023)), which have demonstrated strong results across various tasks such as secondary structure prediction, protein cellular localization or contact prediction. ESM2 has also been evaluated, with the ESM2 3B and ESM2 650M versions (Lin et al., 2023). ESM2 has shown competitive performance with AlphaFold2 in protein structure prediction (Manfredi et al., 2025), indicating that these PLMs have learned important structural information that may be useful in pathogenicity prediction. Additionally, our analysis includes the latest PLM from the ESM Team, called ESM Cambrian (ESM C), which is supposed to outperform ESM2 notably in contact prediction (see https://evolutionaryscale.ai/blog/esm-cambrian, access 10/01/2025).

The combination of VEPs has been shown to be a significant improvement in the prediction of mutation pathogenicity, i.e., MetaRNN (C. Li et al., 2022) or BayesDel (Feng, 2017) are often among the top performers in recent benchmarks (Livesey & Marsh, 2023; Radjasandirane et al., 2025). To further improve performance, we propose here to apply this approach directly onto PLMs. Based on this postulate and on our extensive analysis of PLMs, a novel VEP named PATHOS (PATHOgenicity Variant **S**coring), was proposed. It integrates the most powerful PLMs for pathogenicity prediction. Its performance is enhanced by incorporating both conventional features, like allele frequency, and less common features, such as mutation probabilities derived from phylogenetic analyses, increasing overall performance.

This study first shows that for predicting pathogenicity, fine-tune PLMs may not be the best option as they do not improve performance compared with non-fine-tuned PLMs. In contrast, the combination of non-fine-tuned PLMs and added features significantly increased performances, with PATHOS outperforming 65 other VEPs on three clinical datasets used in our previous benchmark (Radjasandirane et al., 2025). Our results also demonstrate, using practical examples, that PATHOS outperforms AlphaMissense in identifying more biologically relevant regions and disease-causing mutations.

Precomputed predictions for all possible missense mutations for all proteins in UniProtKB/Swiss-Prot (The UniProt Consortium, 2025), representing 140 millions mutations are provided to the scientific community on the web server of PATHOS: https://dsimb.inserm.fr/PATHOS.

## Materials & Methods

### 1. Training and validation datasets

Mutation data were retrieved from two famous public databases: ClinVar (https://www.ncbi.nlm.nih.gov/clinvar/, access date: 2023/12/28) (Landrum et al., 2014) and UniProt (https://www.uniprot.org/, access date: 2023/12/29) (The UniProt Consortium, 2025). ClinVar is a public database that sheds light on the relationships between human genetic variants and diseases, providing clinical annotations of pathogenicity. ClinVar variants were selected using the following filters: “Benign”, “Likely benign”, “Likely pathogenic”, “Pathogenic”, “Missense” and “Single nucleotide” terms. Selected variants have only one amino acid change in the protein sequence. Any variants that are classified as “Uncertain significance” or “Conflicting classifications” have been removed to keep an unambiguous dataset. Only data released before May 1^st^ 2021 were kept.

UniProt is a renowned curated protein sequence and functional annotation database. It contains detailed information about protein sequences, structures, and functions, and links human proteins to their involvement in diseases with associated mutations (when available). Known and verified mutations on these proteins have been listed in one file named “humsavar.txt” available in UniProt. For both databases, labels “Likely benign” are considered as “Benign” and labels “Likely pathogenic” are considered as “Pathogenic”.

### 2. Test dataset

Three test datasets have been used. The first dataset is composed of variants manually and clinically annotated from the literature (Gunning et al., 2021). These variants were curated by experts, ensuring reliable and high-quality annotations. We further removed variants suspected to harbor biases such as type I data circularity (Radjasandirane et al., 2025). The second test dataset corresponds to ClinVar variants released after May 1st, 2021. As extensively described in our previous work (Radjasandirane et al., 2025), this filter on ClinVar dataset also reduces the possibility to have data overlap (type I data circularity) with the training set of VEPs used in this study to have the least biases possible in the evaluation. A filter has also been applied to remove more subtle bias from both test datasets (referred as type II data circularity in (Grimm et al., 2015)), which concerns genes having a disproportionate annotation of one class (Benign or Pathogenic).

Finally, the third dataset, named ClinVar_HQ, is a subset of the ClinVar dataset and only integrates high quality variants that have been submitted multiple times and/or reviewed by an expert panel.

### 3. Protein Language Models

Nine PLMs have been selected to predict pathogenicity. Five PLMs come from the Rost Lab, namely ProtT5 (Elnaggar et al., 2022), ProstT5 (Heinzinger et al., 2023), Ankh Base (Elnaggar et al., 2023), Ankh Large (Elnaggar et al., 2023) and the recent Ankh 2 Large (https://huggingface.co/ElnaggarLab/ankh2-ext2), all available on Hugging Face (https://huggingface.co). These PLMs are known for their good performance across various protein tasks by using a T5-based architecture (Raffel et al., 2023). Two other PLMs from the ESM family (from Meta©), specifically ESM2 3B and ESM2 650M were tested.

Recently, the ESM Team has released a new PLM named ESM Cambrian (ESM C) available in three versions: 300M, 600M and 6B parameters (see https://evolutionaryscale.ai/blog/esm-cambrian, access 10/01/2025). They report that the 600M version outperforms ESM2 3B with a significant gain in computational cost and with five fold less parameters to optimize. Unfortunately, the 6B model is accessible only through a token based request system, for which we have not granted access. Therefore, only ESM C 300M and 600M have been used.

ESM C models was downloaded from the HuggingFace repo called Synthyra/ESMplusplus_small and Synthyra/ESMplusplus_large, which are respectively implementations of ESM C 300M and ESM C 600M allowing inference by batch, since the official ESM C can be used with only one sequence at a time. See Table S1 for specific versions used for each PLM.

### 4. Model architecture

For each PLM, a simple model made of one neuron layer is built, taking as input embeddings from the PLM. An embedding is a vector which implicitly represents information such as the structural environment of the position and physico-chemical properties of the encoded residue. These embeddings, combined with additional features, constitute the features vector that will be fed into the fully connected layer to predict pathogenicity for one PLM (see Figure 1A).

**Figure 1:**
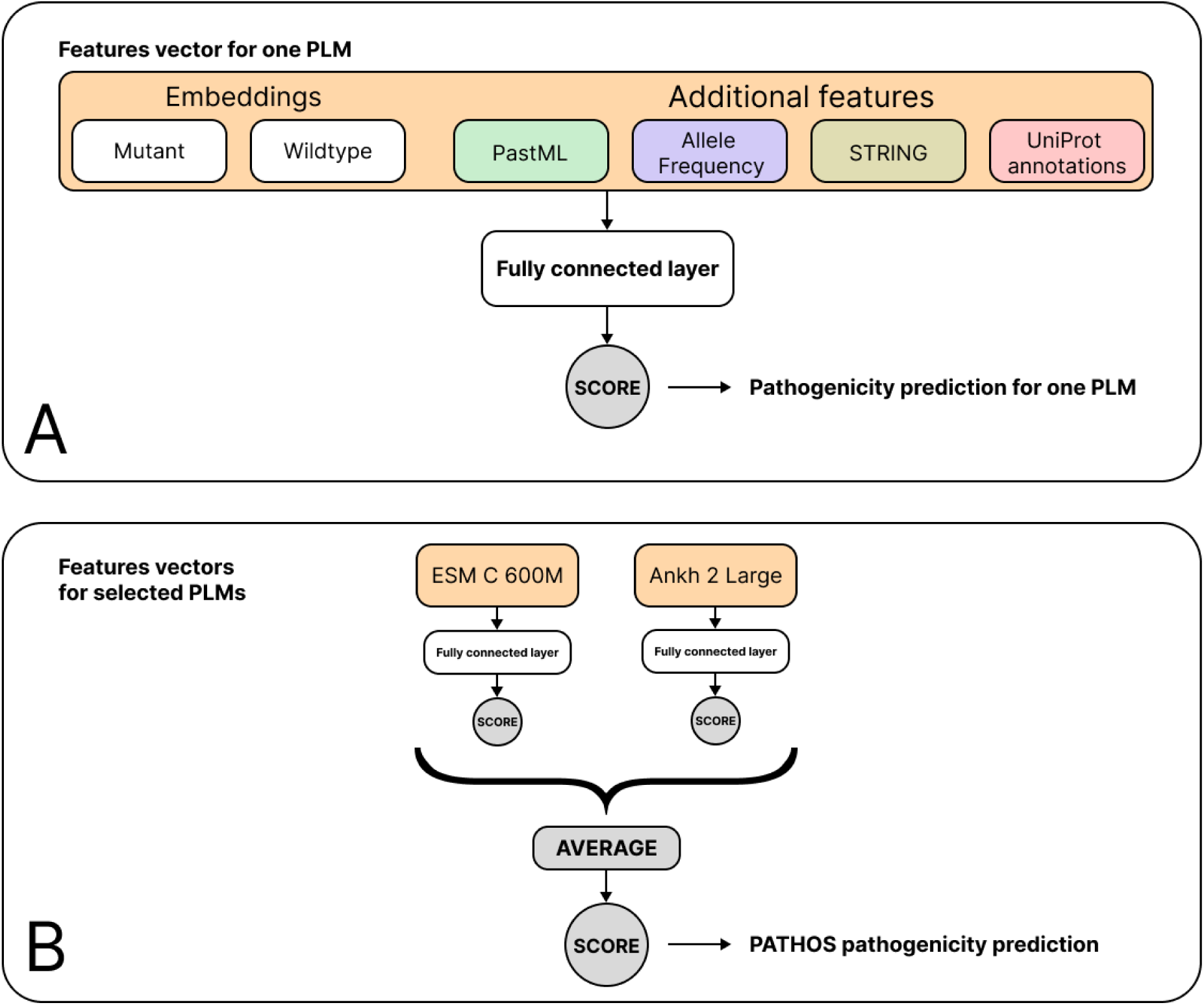
PATHOS framework. **A.** Architecture for one PLM to predict pathogenicity score using a feature vector. This vector is composed of embeddings of wild-type and mutant sequence, and additional features such as PastML probability, allele frequency, STRING and UniProt annotations. This vector is fed into a fully connected layer to predict the pathogenicity from one PLM. **B.** The final PATHOS framework, corresponds to the combination of features vectors from two selected PLMs: ESM C 600M and Ankh 2 Large. Each feature vector is processed through its own fully connected layer to generate an individual score. These two scores are then averaged to obtain a final score which corresponds to the PATHOS pathogenicity prediction.

The combination of the nine PLMs have been tested and our final model PATHOS is an optimal combination of two PLMs, namely ESM C 600M and Ankh 2 Large (see Figure 1B). The predicted scores for each of these two PLMs are averaged to get the final prediction score.

### 5. Features incorporation

Each section here describes a feature that is part of the global vector used to predict pathogenicity in our models (see Table S2).

#### 5.1. PLMs embeddings

For each mutation in our datasets, the mutant sequence was embedded residue by residue, and the embedding corresponding to the mutated position was extracted. Nonetheless, relying only on the mutant embedding could limit the model as it has no reference of the initial residue of the sequence. A change from the original residue (aa1) to a new different one (aa2) may have more impact than a change from aa2 to aa1 depending on the sequence and structure context. Consequently, the wildtype embedding was also generated, making the combination of the mutant and wildtype embeddings a major feature for the trained model.

#### 5.2. PastML

PastML infer ancestral character based on rooted phylogenetic trees at residue-level (Ishikawa et al., 2019). To the best of our knowledge, this study is the first to include PastML into a predictor to predict pathogenicity. The main idea is to traceback the evolutionary history of the mutated position to see if the mutated residue has already been observed in another organism, given residues at this position in homologs sequences. PastML’s equations (Ishikawa et al., 2019) show that the likelihood is influenced by evolutionary distance. A mutation observed closer to the human reference (at a short phylogenetic distance) will contribute more to a high likelihood, meaning a low probability that the considered mutation is pathogenic for humans. An opposite reasoning is applied for a mutation occurring in a distant ancestor (or absent from the tree). PastML requires as input the mutated residue from the human sequence and residues at the same position from aligned homologous sequences in order to construct the phylogenetic tree. Homologs were searched using MMseqs2 (Steinegger & Söding, 2017) against a locally built mammalian database. This search was parametered to allow a maximum of 5,000 sequences sharing at least 50% of sequence identity with the query sequence. Mammalian sequences were extracted from UniProt (accessed on: 02/08/2024) using the taxonomy search. It is composed of 6,435,226 sequences from 2,600 unique species. The query sequence and its homologs are aligned to get the Multiple Sequence Alignment (MSA) with MMseqs2. Finally, the input file required by PastML is built by specifying the mutated residue for the human sequence and the observed residue for all other sequences in the MSA. PastML was run with default parameters and extracted the probability of observing the given mutation computed using marginal posterior probabilities approximation in a Jukes Cantor model (Jukes & Cantor, 1969) (see PastML documentation at: https://pastml.pasteur.fr/help, accessed on: 15/08/2024).

#### 5.3. Allele Frequency

Allele Frequency (AF) information is an important feature as it has been shown to significantly increase predictive performance for pathogenicity (Alirezaie et al., 2018; C. Li et al., 2022). AFs come from the recognized gnomAD v4.1 database (S. Chen et al., 2024), compiling a total of 27,356,284 millions of frequencies available. AF values of variants absent from gnomAD were assigned to zero, as this value is an acceptable approximation for unseen variants.

#### 5.4. String

To feed the model about the interaction propensity of the protein, an interaction score based on the STRING database is added (Szklarczyk et al., 2025). STRING interactions data with a lower quality score than 500 have been removed. The interaction score of each protein was calculated as the division of its number of interactions by the maximum number of interactions observed in STRING (held by TP53 with 1584 interactions). It provides a STRING-based score of the interaction occurrence per protein. Some proteins do not have a STRING entry. For these proteins, a STRING value is assigned, corresponding to the average of the normalized STRING value from the training set (see normalization section below).

#### 5.5. UniProt

UniProt annotations at residue-level such as presence of glycosylation, binding site or secondary structure information have been incorporated into the model. The local information held by neighbors residues of the mutated position within a window of 11 residues centered on the mutated position (+ or − 5 residues around the position) is used as a feature. If the mutation occurs at one of the extremities of the sequence, the window is adjusted to maintain a total of 11 residues. Each variant has a one-hot encoded vector of 34 values (one value per annotation), indicating the presence (1) or absence (0) of specific annotations (see Table S2 for full list of annotations) on the mutation position and among residues surrounding it. After a feature importance analysis, the final PATHOS model incorporated only 18 of these annotations for each variant.

#### 5.6. Features normalization

Features have been normalized through log2 minmax normalization as follows:

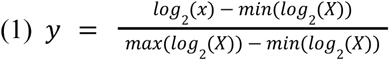

It provides PastML, AF and STRING features with a more homogenous distribution across variants, as they mostly follow extreme law values.

### 6. Training

All training were performed on the HPC resources of IDRIS (Jean-Zay machines) with one A100 GPU per training (Allocation number: AD011014139).

Fine-tuning was tested for each PLMs except ESMC models. Low-Rank Adaptation of Large Language Models (LoRA) approach was applied (Hu et al., 2021), it allows modification of only a fraction of parameters of a large model. It ensures a more efficient and accelerated training process avoiding the computational cost of a full model retraining and potentially a catastrophic forgetting of the initial knowledge of the PLM. A final fully connected layer processes the modified embedding to predict the variant class. The code released by Rost lab (Schmirler et al., 2024) was adapted to fine-tune PLMs with the LoRA approach (with default parameters from the code with an Adam optimizer, a learning rate of 3e-4 and a batch size of 8). The generation of the ESMC 600M and Ankh2 Large embeddings for 140 million mutations of the human proteome was carried out using several H100 GPUs from the Jean Zay supercomputer (Allocation number: AD010316397).

### 7. Threshold optimization

The optimal threshold for classifying variants based on an averaged score resulting from a combination of PLMs has been determined using the validation dataset (as done in section 2.d of results). First, the validation set is subsampled to create a subset containing 80% of benign variants and 20% of pathogenic variants, based on real-world proportions of variants class. These proportions can be observed in public clinical databases such as ClinVar. Then, the best threshold is determined on this subset that gives the highest MCC. These two steps are repeated 1,000 times to generate 1,000 subsets and optimal thresholds. Then, the final optimal threshold is the averaged of the 1,000 optimal thresholds.

### 8. Predictors comparison

PATHOS was evaluated against 65 VEPs, some of which are widely used VEPs such as AlphaMissense (Cheng et al., 2023), MetaRNN (C. Li et al., 2022), SIFT (Ng & Henikoff, 2003) or PolyPhen2 (Adzhubei et al., 2010, 2013). All information in the prediction retrieval and other information related to analyzed VEPs are described in our previous work (Radjasandirane et al., 2025). See Table S3 for information for each VEP.

### 9. Metrics

A confusion matrix is generated between true and predicted annotations for each VEP that count the amount of variant correctly predicted in each class (benign or pathogenic). From this matrix, the amount of: (i) pathogenic variants predicted as pathogenic (True Positive or TP), (ii) benign variants predicted as benign (True Negative or TN), (iii) pathogenic variants predicted as benign (False Negative or FN) and (iv) benign variants predicted as pathogenic (False Positive or FP) can be extracted. This confusion matrix is used to compute sensitivity, specificity, Balanced Error Rate (BER), the Matthews correlation coefficient (MCC) and F1 score metrics with following classical formulas:

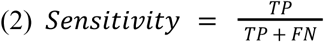

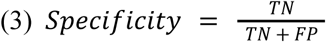

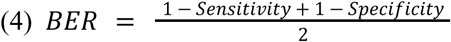

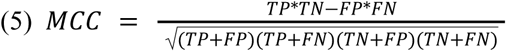

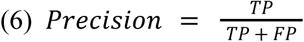

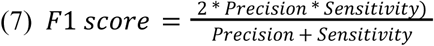

For each dataset and metric, the statistical significance of PATHOS performance has been assessed using a bootstrap method on 10,000 iterations. PATHOS performance was compared against the top ranking VEPs: the four best performing VEPs (excluding PATHOS) if PATHOS is already among the top five, or the five best performing VEPs otherwise.

The rank score metric was used as the last main metric, which has been introduced by (Livesey & Marsh, 2023), and adapted in our benchmark (Radjasandirane et al., 2025). This score evaluates a pair of VEPs only using shared variants between these two VEPs, ensuring a fair comparison between them. The VEP having the highest MCC accumulates one winning point if the difference is statistically significant (based on a McNemar test), or 0.5 if they both have similar predictions distribution. The final rank score for a given VEP is the mean of its win and draw results. As the calculation of the rank score incorporates a statistical test, no additional significance computations were performed, unlike with classical metrics.

## Results

### 1. Datasets

From the ClinVar database, 71,752 variants were extracted, including 38,504 labeled benign and 33,248 pathogenic. The Humsavar dataset contains 72,585 variants, including 39,755 benign variants and 32,805 pathogenic variants. Concatenating the two sets resulted in a dataset containing 128,342 variants, with 73,130 benign and 55,212 pathogenic variants (15,995 duplicate variants were removed). Variants whose sequences did not match the canonical UniProt sequence (version 2025_01, downloaded on 03/20/2025) were removed, accounting for 16,566 variants. The remaining 111,768 variants underwent two quality filters.

The first filter addresses over-represented genes. Data from clinical context indeed tends to over-represent genes most frequently encountered in the clinical setting. In this case, some genes in the dataset have over 400 variants, including the FBN1 gene, which possesses 1038 variants by itself. We therefore removed the top 1% of the most represented genes, which was sufficient to ensure that the most represented remaining gene had at most 90 variants. This filter removed 136 genes and 23,028 variants.

The second filter concerns the type 2 data circularity bias which occurs when genes show a strong dominance for a specific annotation, such as “pathogenic”. A model trained on such genes would risk assimilating this strong correlation, thus learning that the gene is very often associated with pathogenic variants. This bias is then likely to affect evaluations by artificially inflating the performance of the model. Consequently, we removed the genes that were imbalanced. In our previous work (Radjasandirane et al., 2025), we chose thresholds of 40% and 60% to define a balanced gene. However, using these thresholds on our dataset would remove far too much data (90%), making the training unstable. We opted for less strict thresholds: a gene is considered balanced if the proportion of its annotations falls between 20% and 80%. To calculate representative proportions, genes must have at least 5 variants; otherwise, they are removed. This second filter removes 70% of the variants that passed the first filter.

These two filters thus removed 11,894 genes (representing 88% of the initial genes) and 85,201 variants (representing 76% of the initial dataset), leading to a final dataset composed of 26,566 variants, including 13,984 benign variants and 13,582 pathogenic variants. This data processing removes potential biases inherent in clinical annotation data, thereby making the model training much more robust.

To construct the training and validation datasets, a protein-based split was performed to avoid any overlap and additional bias between the two sets. Consequently, 80% of the unique proteins were assigned to the training set, and the remaining 20% to the validation set. This approach resulted in a training set composed of 21,037 variants and a validation set of 5,529 variants, with a balanced class distribution in both sets.

### 2. Model development

#### a. Impact of fine-tuning on PLMs performance on detecting pathogenicity

LoRA based fine-tuning has been shown to improve PLM performances on some tasks (Schmirler et al., 2024); it was so evaluated to assess its impact on variants pathogenicity prediction. Seven PLMs have been fine-tuned and their performance was compared to the original versions of these PLMs. Training was limited to a single epoch as further training reduced performance (data not shown). The performance of each fine-tuned model is measured on the validation dataset using the MCC metric and compared to the MCC of the non-fine-tuned model. Fine-tuning appears to slightly improve performances of ProstT5 models with an increase from 0.517 to 0.533 MCC (see Figure 2A). However, for other PLMs, no strong improvement is observed as MCC values are quite similar between two conditions. Surprisingly, for ESM2 3B, Ankh Large 2 and Ankh Large models, the fine-tuning process is decreasing slightly their performance with respectively a loss of 0.019, 0.049 and 0.027 of MCC. Consequently, it appears that fine-tuning PLMs does not improve performance sufficiently to make it worthwhile in the context of pathogenicity prediction, for the PLMs we consider in our study.

**Figure 2:**
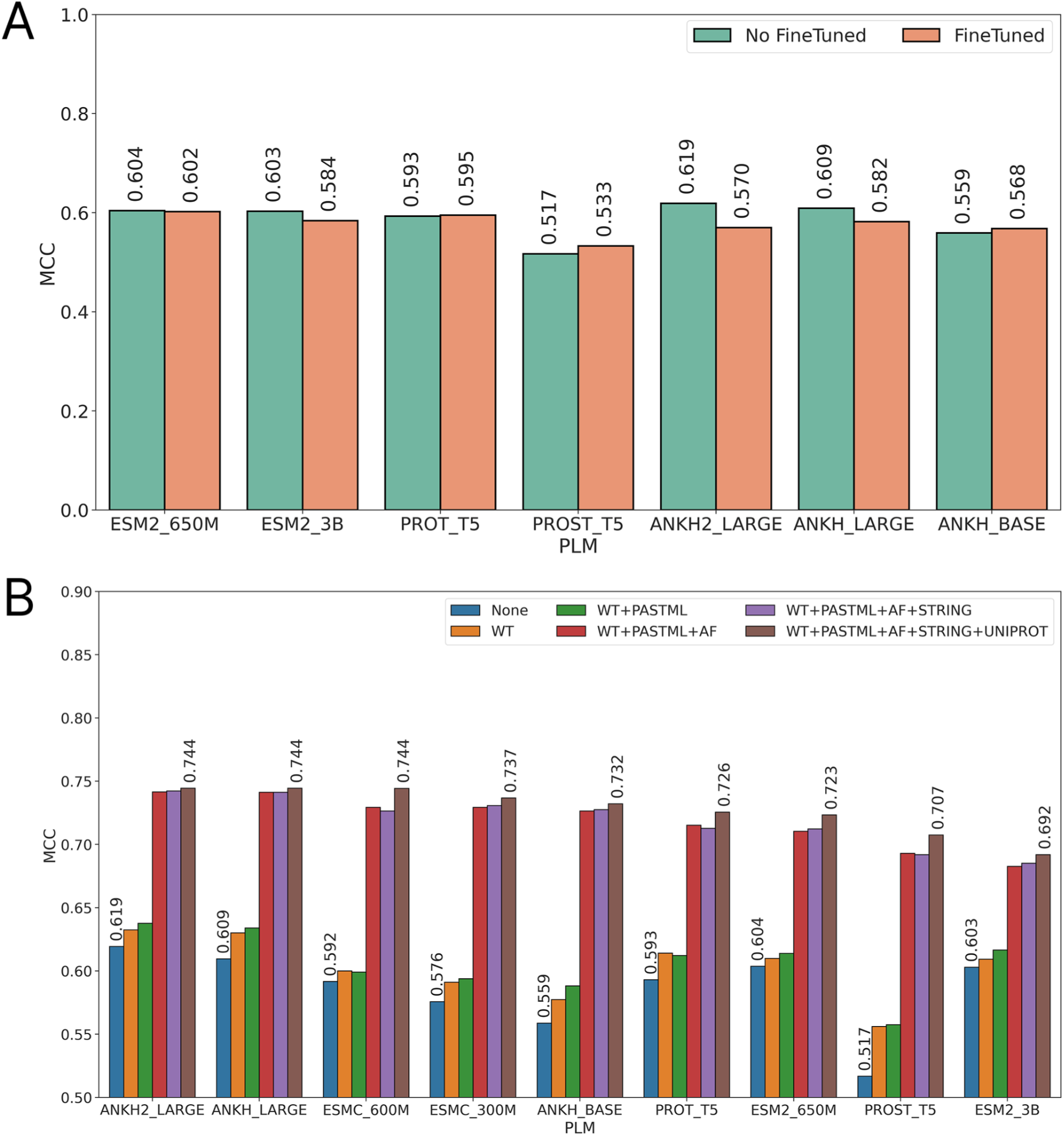
Performance of each PLM computed on validation dataset. (A) Comparison of performance in MCC using embeddings from fine-tuned PLMs (in orange) versus original PLMs (in green) to predict pathogenicity. (B) Performance in MCC using original embeddings only (labelled None) or with additional features such as wild-type embeddings (WT), PastML probability (PASTML), Allele frequency (AF), STRING score (STRING) and UniProt annotation vector (UNIPROT).

Furthermore, fine-tuning significantly affects computational time (see Table S4). Indeed, training times are relatively high as the model generates embedding on the fly for all mutations and for each batch of data, as the embeddings are modified alongside with the weights optimization. The repetition of embedding generation, training computations and weights optimization steps can be inefficient. Surprisingly, fine-tuning ESM2 3B is as fast as fine-tuning Ankh base model despite the important difference in the number of parameters. This has been discussed in (Schmirler et al., 2024), LoRA seems to be more efficient for larger models. On the other hand, the non-fine-tuning process is more efficient as embeddings can be pre computed before the training and can be reused for all different experiments, which reduces both training time and implementation complexity. An homemade API script (available at https://gitlab.dsimb.inserm.fr/cretin/llm-api) that handles batches of sequences efficiently and asynchronously has been used, which significantly accelerates embedding generation. Additionally, since PLMs are publicly accessible on HuggingFace, it is more convenient for the community to use them to generate original embeddings. Therefore and given the negligible performance gains from fine-tuning, we decided to use original (non-fine-tuned) PLMs for all further analyses.

#### b. Incorporation of biological features to improve performance

Adding features greatly improves the overall performance of each PLM (see Figure 2B). Among these features, AF radically changes the understanding of the model by increasing the MCC by more than 0.1 for almost all PLMs, bringing them above 0.70 of MCC. The main drawback of using AF as a feature is the important proportion of missing data in gnomAD. Indeed, roughly 35% of the train and validation datasets have unavailable AF. The second best feature is the wild-type embeddings as it informs each model the initial sequence state before mutation, which is important to correctly understand the impact of the mutation on the protein. PASTML, STRING and UniProt features further increase performance of all PLMs.

With all features, the best-performing PLMs are Ankh2 Large, Ankh Large, and ESMC 600M, reaching an MCC of 0.744. The lighter models of these PLMs achieve slightly lower performance, with 0.737 for ESMC 300M and 0.732 for Ankh Base. It is interesting to note that ESM2 3B offers the lowest performance, with an MCC of 0.692 when all features are added. When the model uses only the information from the mutant embedding, the MCC is 0.603, corresponding to the 4th best MCC (without added features, in blue in Figure 9.4B). Despite the large difference in model size, ESM2 650M achieves better performance than its three-billion-parameter version with an MCC of 0.723, highlighting good compatibility of ESM2 650M with the added features. As indicated by the authors of ESMC, our results confirm that ESMC 600M, and even the 300M version, do indeed seem to surpass ESM2 3B for this task (Figure 2B) with a much lower number of parameters (Table S4). ESMC stands out as an accurate and efficient PLM to integrate into variant pathogenicity prediction methodologies.

The UniProt feature vector is initially composed of 34 annotations extracted from the given protein’s UniProt entry (Table S4). The utility of each annotation was evaluated using the SHAP methodology (Lundberg & Lee, 2017), which estimates the importance of each feature integrated into a model. This analysis highlights the relevance of annotations related to secondary structure, such as ‘Beta strand’, ‘Helix’, or ‘Topological domain’ (Figure S1A), which are among the four most useful annotations. The presence of known variants in the area of the mutated position also impacts the model’s predictive capacity via the ‘Natural variant’ annotation, which is the third-best annotation. Other annotations, such as ‘Turn’, ‘Transmembrane’, or ‘Disulfide bond’, have relatively lesser importance but still impact the prediction. Finally, some features have a SHAP value close to zero, such as ‘Chain’ or ‘Lipidation’, indicating that the annotation is either not useful for the predictive task or that the quantity of variants for these cases is too low. To remove unnecessary annotations, the cumulative sum of SHAP values was calculated. The retained annotations are those sufficient to explain 95% of the UniProt features (Figure S1B). This process reduced the length of the UniProt feature vector from 34 to 18 annotations, leading to the removal of annotations such as ‘Motif’, ‘Chain’, or ‘Peptide’ (see Table S2).

#### c. Analysing redundancy between PLMs

The different PLMs have been trained differently with distinct training datasets (Elnaggar et al., 2022; Rives et al., 2021). Each has incorporated unique knowledge about amino acids from their training. To assess whether the PLMs predicted pathogenicity in a similar manner, the percentage of identical predictions for each PLM pair was calculated. Within these predictions, a second percentage indicating the correct predictions was also determined (see Figure S2). Ankh Large and Ankh2 Large show a very high correlation of 96%, which is expected since Ankh2 Large is a more optimized version of the same model. Another expected result is the strong correlation between ESMC 300M and ESMC 600M, which have a 90% correlation.

Interestingly, the Ankh models show a better correlation with the ProtT5 and ESMC models, with correlations between 87% and 88%, but are slightly less correlated with the ESM2 models, with correlations around 86%. Similar observations can be made with the ESMC models compared to the Ankh, ProtT5, and ESM2 models. The percentage of identical correct predictions barely exceeds 74% for all pairs. Moreover, the correlations indicate that the level of agreement varies between the different PLM pairs. There are thus notable divergences in their predictions, as no correlation is close to 100%. An optimal combination of these PLMs would have the potential to create a more robust model, where each PLM might be able to compensate for the errors of the others.

#### d. Find an optimal combination of PLMs

We hypothesize that a unified tool built from an optimal combination of PLMs can significantly improve performance. To find this combination, each PLM is first evaluated individually, using its initial MCC as a baseline to improve. For each of these starting PLM, one candidate PLM at a time is added iteratively. For each new combination, the average score across the current set of selected PLMs is computed. Based on this averaged score, the best possible threshold to maximize the MCC is determined (as described in section 7 of Materials and Methods). If adding the candidate PLM improves the MCC over the current combination, the PLM is retained (marked as green, see Figure 3) and the resulting MCC becomes the new baseline to improve. Otherwise, the PLM is discarded (marked as red).

**Figure 3:**
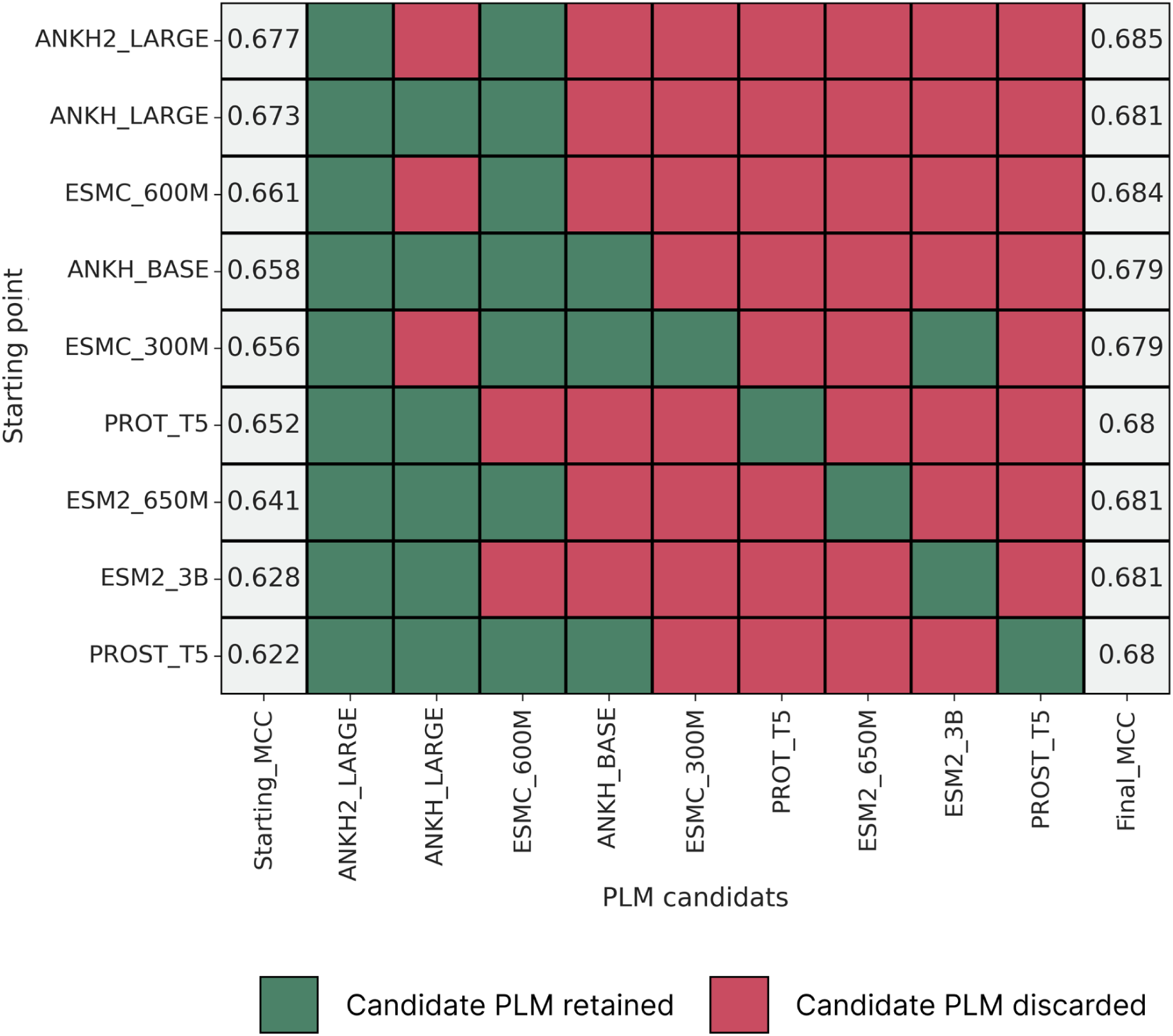
Combination analysis of PLMs. The initial PLM used is shown in the y-axis. The first column in the x-axis is the starting MCC for each PLM that needs to be improved through adding scores of other PLMs. If adding the candidate PLM increases the actual MCC of the combination, it is retained and marked in green, otherwise, it is not retained and marked in red. For each PLM, all PLMs highlighted in green constitute the final and optimized combination, and the corresponding final MCC for that combination is shown in the last column.

This process is repeated for each PLM candidate to test. This approach ensures that only models that increase performance are included, resulting in a combined predictor that outperforms any single PLM alone. The starting MCCs differ from those illustrated in Figure 2B for the validation set because here, they are calculated using an optimized threshold. This threshold was specifically determined by balancing the class distribution with an 80% benign and 20% pathogenic variants proportion (selected randomly), which leads to a different MCC value (see section 7 of Materials and Methods).

Following this strategy, the best combination is Ankh2 Large combined with ESMC 600M, yielding an MCC of 0.685 (0.684 when starting from ESMC 600M). Despite the small gap between the final MCC values of the combinations, this solution is the most parsimonious as it only uses two PLMs, unlike other solutions with 3 to 5 PLMs, while also achieving a high MCC. For this specific combination, an optimal threshold of 0.63 was determined using the iterative process described in section 7 of Materials and Methods. The combination of these two PLMs will be designated as PATHOS in the remainder of the text.

### 3. Comparing PATHOS to other VEPs

PATHOS was evaluated on clinical variants using three main metrics: MCC, F1-score, and rank score. It should be noted that recent UniProt updates have modified some canonical sequences. The sequences of mutants in the test datasets may no longer match, as these datasets were built with an older version of UniProt (old UniProt version used for datasets construction: 2023_01, version used here: 2025_01). Consequently, a very small number of variants (less than 60 in total) do not have a PATHOS score because they no longer correspond to the canonical sequence of the UniProt version previously used to build the test datasets (Radjasandirane et al., 2025).

#### a. PATHOS achieves state of the art performance on manually annotated variants

On the Clinical dataset, PATHOS achieves a MCC value of 0.591, statistically outperforming by a large margin top VEPs on this dataset, including gMVP which is second with a MCC of 0.508 (see Figure 4A). This increased MCC value indicates that PATHOS has better prediction on both benign and pathogenic classes compared to other VEPs. PATHOS ranks first for the F1 scores with a score of 0.785, beating VARITY_R which has a F1 score of 0.744 (see Figure 4B). This highlights PATHOS ability to accurately identify true positives while minimising false positives and false negatives. Clinical applications are heavily dependent on this point especially when searching for pathogenic variants among a patient genome that harbors a large fraction of benign variants.

**Figure 4:**
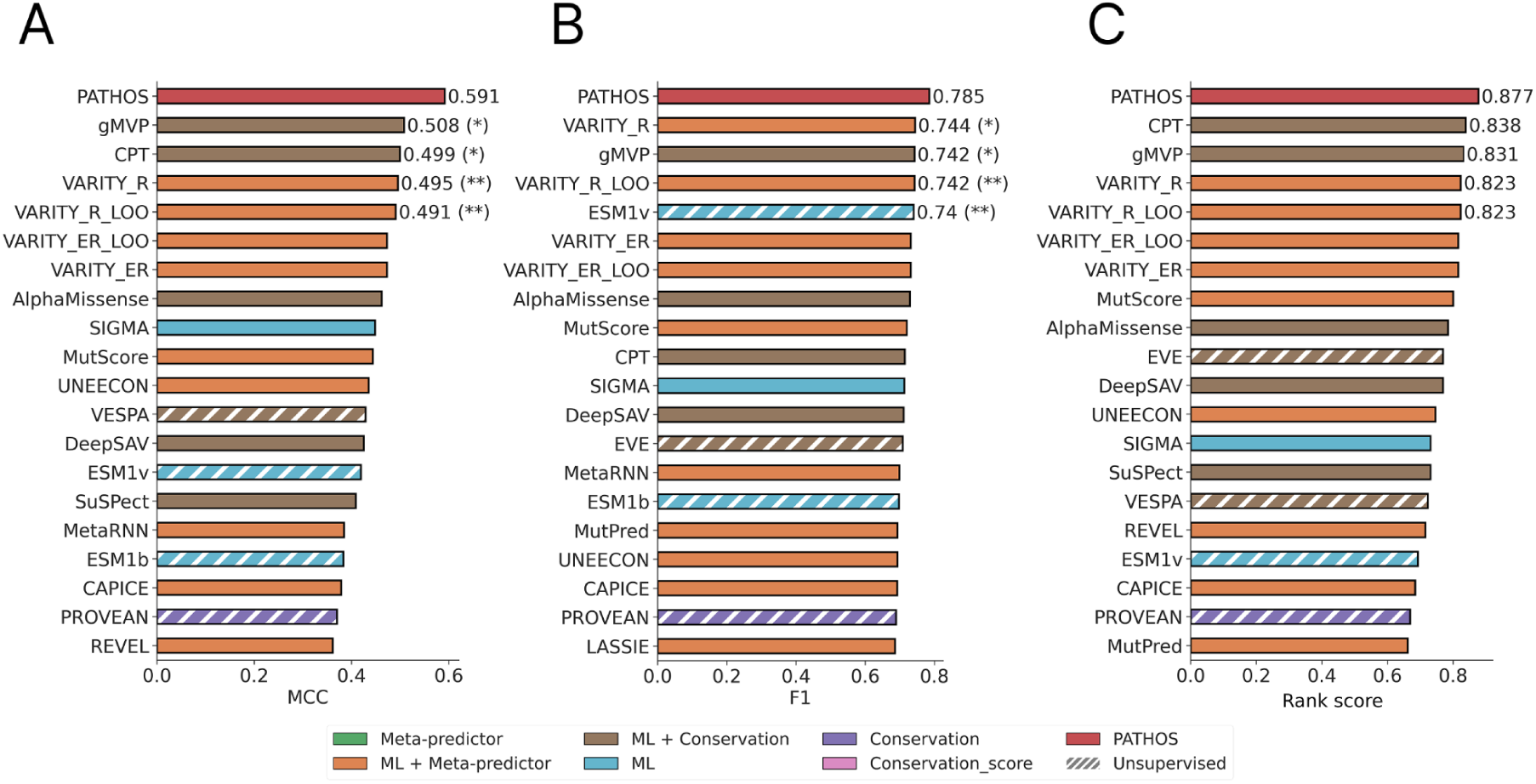
Performance on Clinical dataset. Histogram of MCC (A), F1 (B) and Rank score (C) values for the Clinical dataset. Only the top 20 VEPs out of 66 are shown. For the performance of all 66 VEPs on Clinical datasets, refer to Figure S3 A, B, and C. Each VEP is colored based on its algorithm type used as depicted by the legend on the bottom of the figure. Statistical differences between PATHOS and top-ranked VEPs have been assessed using the bootstrap method on 10,000 iterations.

Finally, PATHOS achieves the best rank score with a value of 0.877 (see Figure 4C), meaning that it performs better or equally than 87.7% of 65 VEPs on this dataset. The second best rank score is detained by CPT with a score of 0.838. These performances on the Clinical dataset are quite convincing knowing that these variants are manually curated from the literature, making the Clinical dataset a high quality one.

#### b. Optimal performances of PATHOS on the public ClinVar database

PATHOS have also been evaluated on ClinVar variants, as it remains an important clinical resource publicly available. The same dataset as in our previous work was used; namely ClinVar and ClinVar_HQ datasets (Radjasandirane et al., 2025).

PATHOS achieves first rank across all metrics on the ClinVar dataset. It performs similarly than MetaRNN, but statistically outperforms BayesDel and ClinPred, which have consistently ranked among the top three VEPs in many benchmarks using ClinVar dataset including ours (Livesey & Marsh, 2023; Radjasandirane et al., 2025). PATHOS reaches a MCC of 0.826 on the ClinVar dataset (see Figure 5A), a F1 score of 0.855 (see Figure 5B) and a Rank score of 0.954 (see Figure 5C), while BayesDel and ClinPred achieve respectively 0.791 and 0.777 for MCC, 0.825 and 0.807 for F1 score and 0.923 and 0.885 for rank score, respectively. Given that the ClinVar dataset is imbalanced toward the negative class, the high F1 score of PATHOS is particularly significant, as there is a higher proportion of benign variants that can be misclassified as pathogenic variants. Despite this imbalance, PATHOS maintains robust performance by correctly identifying pathogenic variants without overpredicting them. On the higher-quality ClinVar dataset (ClinVar_HQ), PATHOS ranks second on the MCC and F1-score metrics but with no significant difference from the top VEPs, with an MCC of 0.836 (Figure 5D) and an F1-score of 0.889 (Figure 5E). MetaRNN takes first place with an MCC of 0.859 and an F1-score of 0.906. PATHOS achieves a rank score of 0.8, beating the previous top VEP on this dataset, namely the popular AlphaMissense, which has a rank score of 0.762 (Figure 5F).

**Figure 5:**
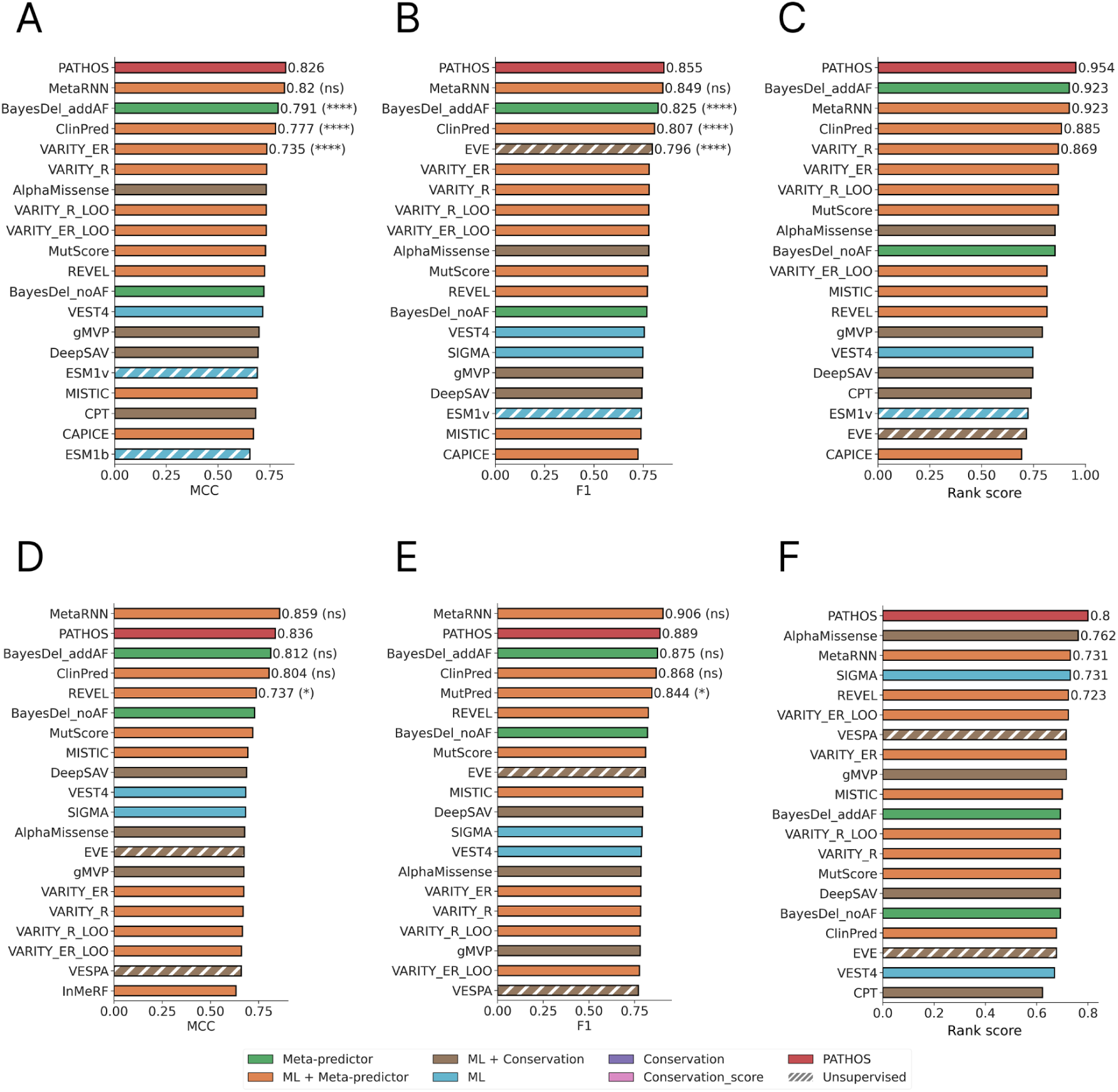
Performance on ClinVar and ClinVar_HQ dataset. Histogram of MCC (A), F1 (B) and Rank score (C) values for the ClinVar datasets, and MCC (D), F1 (E) and Rank score (F) for the ClinVar_HQ dataset. Only the top 20 VEPs out of 66 are shown. For the performance of all 66 VEPs on ClinVar datasets, refer to Figure S4 A, B, and C. For their performance on ClinVar_HQ, see Figure S5 A, B, and C. Each VEP is colored based on its algorithm type used as depicted by the legend on the bottom of the figure. Statistical differences between PATHOS and top-ranked VEPs have been assessed using the bootstrap method on 10,000 iterations.

#### c. Overall performance of PATHOS

The average rank score is computed using the Clinical and ClinVar_HQ dataset in order to have an overview of the performance of VEPs on all high quality clinical variants. PATHOS achieves an averaged rank score of 0.838 (see Figure S6), beating the previous best VEP on this analysis which was AlphaMissense with a score of 0.773, tied with gMVP. This demonstrates that, overall, PATHOS yields excellent prediction performance on our clinically annotated variants.

#### d. Generalization capability of PATHOS on new sequences

To evaluate if PATHOS can generalize well, its performance was assessed using only new sequences that had not been encountered during training. For each test dataset, mutation data was excluded if the protein was present in the training dataset, remaining only ‘new’ sequences. A significant portion of the evaluation data already corresponds to new sequences (Figure S7), due to our data processing that removed a large number of genes from the initial data pool.

PATHOS demonstrates strong generalization capabilities on new sequences, consistently achieving top performance. On the Clinical dataset, PATHOS reaches an MCC of 0.619 (Figure 6A), an F1-score of 0.809 (Figure 6B), and a rank score of 0.808 (tied with CPT, Figure 6C). gMVP ranks just behind PATHOS for MCC (0.559) and F1-score (0.777), with no significant differences with PATHOS.

**Figure 6:**
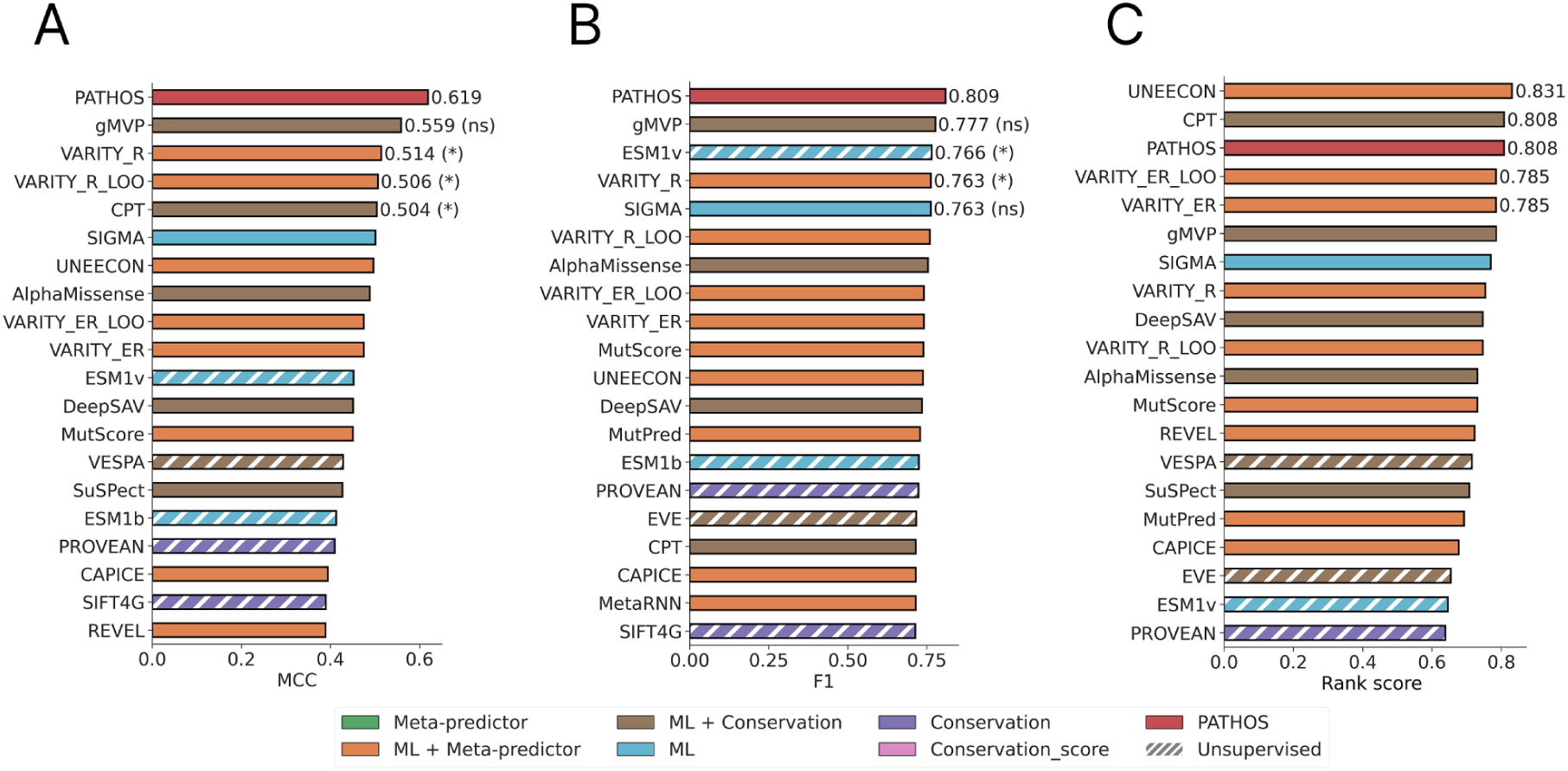
Performance on Clinical dataset using new sequences absent from the training dataset. Histogram of MCC (A), F1 (B) and Rank score (C) values for the Clinical datasets. Only the top 20 VEPs out of 66 are shown. Each VEP is colored based on its algorithm type used as depicted by the legend on the bottom of the figure. Statistical differences between PATHOS and top-ranked VEPs have been assessed using the bootstrap method on 10,000 iterations.

Performances on new ClinVar sequences are relatively similar to the performances calculated on all genes, given that a significant portion of the initial sequences are new. PATHOS achieves first place on all metrics with an MCC of 0.824 (Figure S8A), an F1-score of 0.844 (Figure S8B), and a rank score of 0.931 (Figure S8C). Due to the resemblance to the initial ClinVar dataset, the top VEPs are still MetaRNN, BayesDel, and ClinPred. Except for MetaRNN, these VEPs show statistically different performances compared to PATHOS.

On the ClinVar_HQ dataset, PATHOS is second but without statistical differences, likely due to the reduced size of the dataset (Figure S7). It achieves an MCC of 0.861 (Figure S8D), an F1-score of 0.901 (Figure S8E), and a rank score of 0.8 (Figure S8F). These results suggest that PATHOS can easily generalize to new sequences with high performance. These findings demonstrate that PATHOS training allows it to capture robust features to distinguish pathogenic from benign variants, even on new sequences.

### 4. Performances of PATHOS in detecting Hard variants

We previously showed that variants can be classified into three groups of prediction difficulty: easy, moderate and hard (Radjasandirane et al., 2025). These groups were constructed based on the proportion of error made by VEPs on variants. A variant having less than 30% of errors from VEPs is classified as easy, whereas a variant with more than 70% of error will be in the group hard. Otherwise, the variant is considered as of moderate difficulty.

The performance of PATHOS has been evaluated on all three classes of difficulty against VEPs among the best performing ones in each dataset, namely AlphaMissense, MetaRNN, BayesDel_addAF, ClinPred, VESPA, CPT and gMVP. For each class of difficulty, the performance of VEPs is measured by calculating the MCC metric on the corresponding variant subset. On the ClinVar dataset (see Figure 7A), all VEPs appear to perform similarly on easy variants except for VESPA showing a lower MCC value of 0.773. Performances on moderate variants are more disparate, but PATHOS is better by almost 0.1 from ClinPred (the second best on moderate variants). MCC values for hard variants are negative as these variants exhibit complex characteristics often missed by VEPs. The best VEP for these variants correspond to the least negative MCC, indicating that the considered VEP is less wrong than others. It appears that PATHOS performs relatively well with a MCC of −0.342, Clinpred is not that far with a MCC of −0.355. They both perform better than others on hard variants of ClinVar dataset. Similar observations can be done for the ClinVar_HQ dataset (see Figure 7B), except that PATHOS and ClinPred perform similarly for hard variants with a MCC of −0.107. For the Clinical dataset (see Figure 7C), MCC are lower for all classes especially for moderate and hard variants. PATHOS achieves the best MCC for moderate variants with a MCC of 0.483, where the second best VEP, namely gMVP, reaches only 0.245. PATHOS achieves a MCC of −0.478 on hard variants of the Clinical dataset, behind VESPA and CPT having MCC values of −0.367 and −0.392 respectively. One major observation is that these VEPs are mostly better than AlphaMissense to distinguish easy from hard variants, and that each difficulty class has its own distribution of MCC values, which are almost similar across datasets in terms of VEPs ranking.

**Figure 7:**
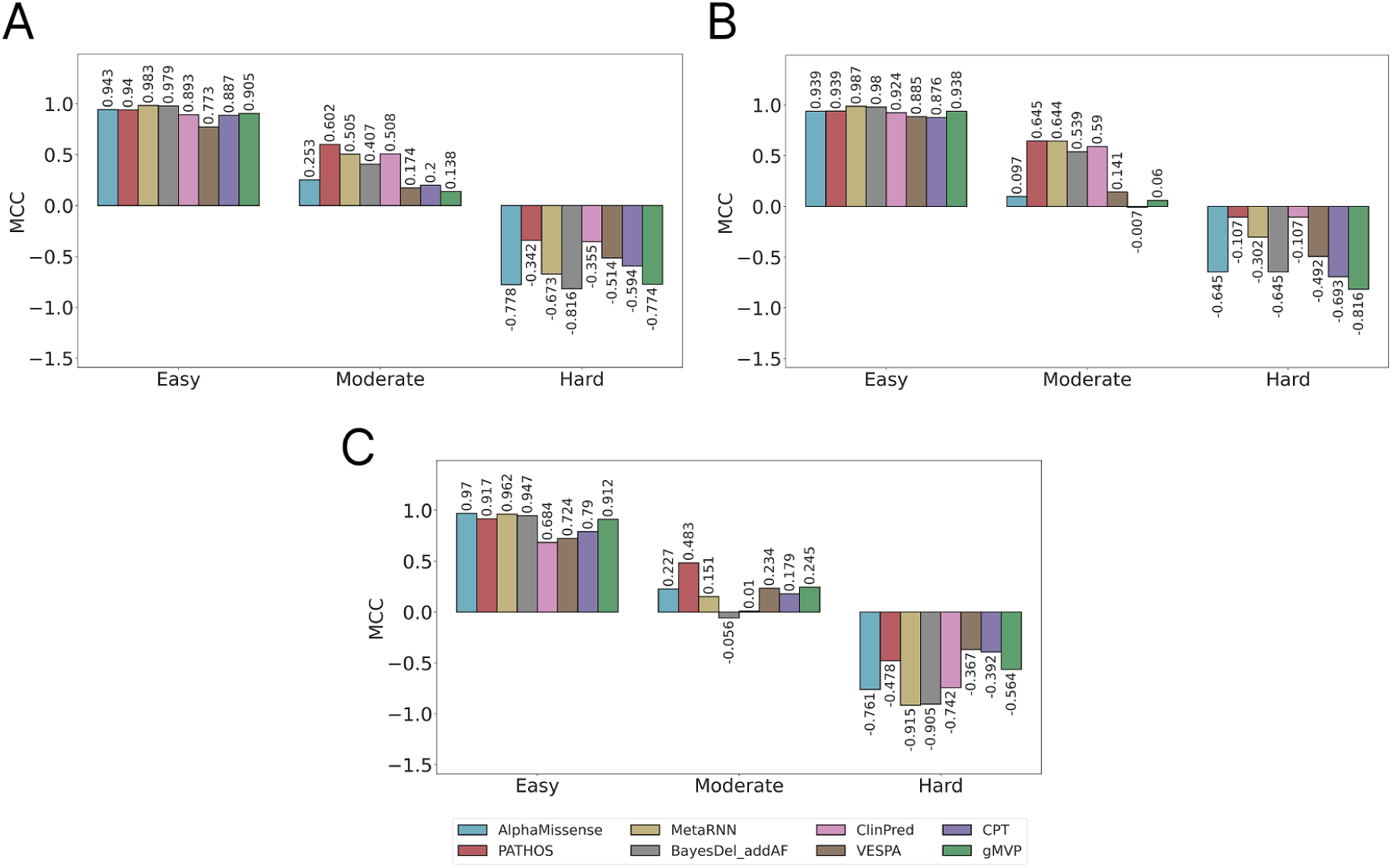
Performance on each variant difficulty class. MCC values for AlphaMissense (in cyan), PATHOS (in red), MetaRNN (in yellow), BayesDel_addAF (in grey), ClinPred (in pink), VESPA (in brown), CPT (in purple) and gMVP (in green) for ClinVar (A), ClinVar_HQ (B) and Clinical (C) datasets.

### 5. Application of PATHOS on the progesterone receptor

One interesting application of VEPs is to predict the impact of all single point mutations possible for a given protein, resulting in a pathogenicity map that can highlight important regions of the protein. This idea has been applied to the progesterone receptor protein (PGR or NR3C3), comparing our pathogenicity distribution to AlphaMissense one (see Figure 8).

**Figure 8:**
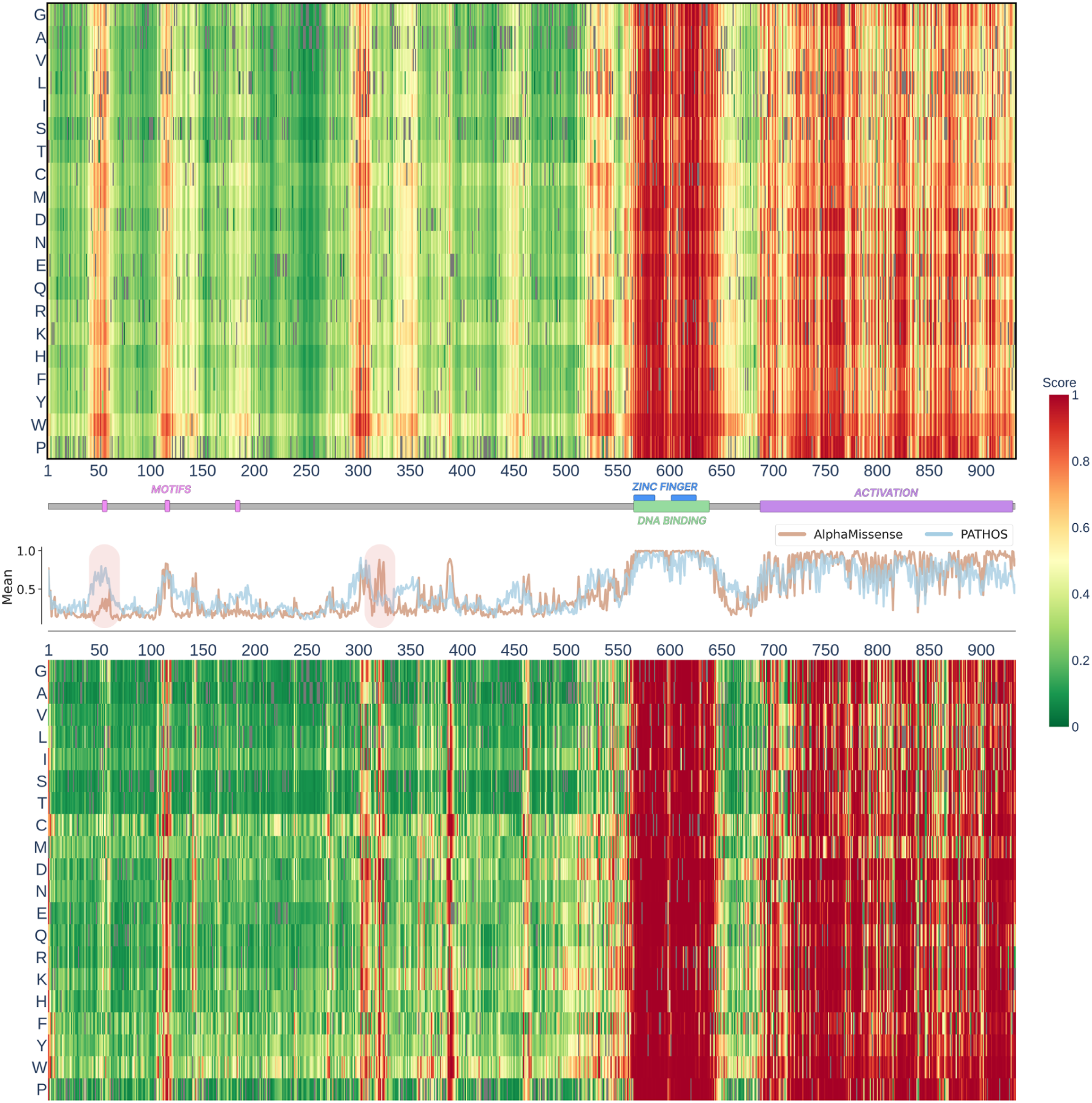
Pathogenicity landscape of the progesterone receptor (PGR, UniProt ID: P06401). From top to bottom: (i) Pathogenicity landscape as predicted by PATHOS. Higher scores indicate higher pathogenicity. (ii) UniProt key annotations for PGR that may correspond to pathogenicity patches. (iii) Plot showing the averaged pathogenicity scores for both PATHOS (in blue) and AlphaMissense (in orange). And (iv), pathogenicity landscape as predicted by AlphaMissense, with the same color scale as PATHOS.

This protein is a hormone receptor that is involved in regulation of gene expression. It has an activator or repressor activity, depending on the expressed isoform. The sequence used corresponds to the activator form and is long of 933 residues. The PGR receptor has many key domains essential for its normal activity, such as its DNA binding region located between residues 567 to 639. A mutation on this regulator can be deleterious by increasing or decreasing the expected activity of the PGR receptor. An example is the mutation Y890C that has been demonstrated to be deleterious by inducing a reduced transactivation capability (Jacobsen & Horwitz, 2012). An impaired PGR receptor by such mutations can lead to breast cancer (Z. Li et al., 2022). Evaluating the impact of mutations on this protein is therefore of high interest.

Compared to AlphaMissense, PATHOS detects the same pathogenicity patches on the sequence, mainly around residue 600 and the big patch from residue 690 to the end of the sequence. The first patch is the DNA binding region of the protein with its two zinc fingers. Any mutation on this important region seems to yield a pathogenic behavior according to both AlphaMissense and PATHOS. The second patch is annotated in UniProt as mediating transcriptional activation, which likely is an important domain for the activity of the receptor. This concordance is also found for the previously mentioned mutation Y890C where both predict this mutation as pathogenic.

In the first half of the sequence, some differences can be noticed between these two distributions. The main differences are around residues 57 and 320 as depicted by the plot of averaged scores by position. While the second region has no particular annotation, the first region is annotated as an important motif in UniProt with an associated article demonstrating the importance of this region (Tung et al., 2001). This motif is known as a LXXL motif, a key motif for the activation of PGR. AlphaMissense is able to detect the second LXXL motif around residue 117, but not the one around residue 57.

PATHOS demonstrates a high sensitivity for functionally important regions within proteins. For example, the model does not predict the motif around residue 185 as an important region, which is consistent with the lack of evidence in UniProt regarding its importance for the receptor. This ability to distinguish between biologically significant motifs and those with lesser functional roles indicates that PATHOS avoids the overprediction of pathogenicity in these regions.

### 6. Investigating the impact of the T587M mutation on the KCNQ1 channel

Ion channels are proteins embedded into the membrane of cells. Their primary role is to allow the selective passage of ions across the lipid bilayer. These channels are key to biological processes from cellular physiology to neuronal communication. An example of an important channel is the potassium voltage-gated channel KCNQ1, its activity is essential for repolarizing the cardiac action potential. This protein is long of 676 residues and is composed of six helices inserted into the membrane of the cell. The fold directly forms the typical channel shape with additional helices, present in the intra cellular environment. The channel interacts with many proteins such as calmodulin or protein kinases and is located in heart muscle cells but also in other tissues such as ear, stomach or kidney (Zheng et al., 2007).

A known mutation that impairs the normal functioning of this protein is the mutation T587M that leads to a Long QT Syndrome (Kapplinger et al., 2009), which is a disease characterized by an abnormal heart electrical activity. Similarly to the analysis of the PGR receptor, the impact of all missense mutations of this protein has been predicted using AlphaMissense and PATHOS. Both mainly detect two pathogenicity patches, the first between residues 110 and 390 (called patch 1 in Figure 9A), and the second between residues 500 and 620 (called patch 2 in Figure 9A). The first patch is the region containing all helices embedded into the membrane and the second one is the intracellular region, interacting notably with the calmodulin as shown in Figure 9C.

**Figure 9:**
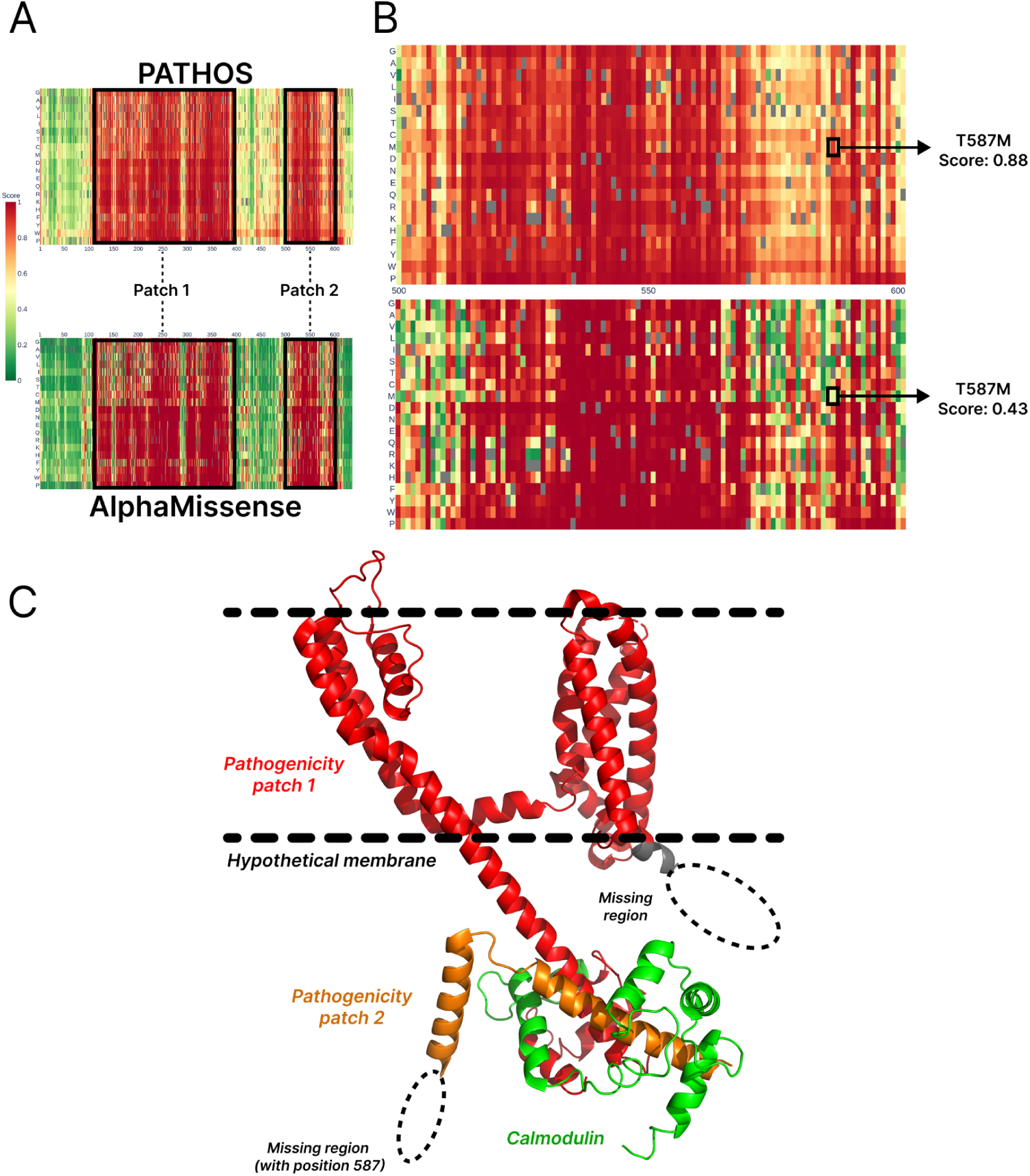
Pathogenicity landscape of the potassium channel (KCNQ1, UniProt ID: P51787). (A) Pathogenicity landscape as predicted by PATHOS (top) and AlphaMissense (bottom). For each heatmap, two regions are highlighted which are the two pathogenicity patches detected by both VEPs, called patch 1 and patch 2. (B) Zoom representation of patch 2, containing the mutation T587M which is highlighted with predicted scores from PATHOS (top) and AlphaMissense (bottom). (C) Structure of the KCNQ1 channel (PDB ID: 7XNN) with an hypothetical membrane position based on annotations from UniProt. Pathogenicity patch 1 observed in (A) is colored in red, and pathogenicity patch 2 in orange. The interacting calmodulin is shown in green. Dashed circles indicate unmodeled regions of the protein (only two of them are represented here), one of them contains the missing position of the mutation T587M.

These pathogenicity patches indicate the high importance of these domains for the normal functioning of the channel. A mutation in the first patch may perturbate the insertion of the channel in the membrane, while a mutation in the second patch can eventually prohibit protein interaction in the intracellular environment. The mutation T587M is located in a coiled coil region (according to UniProt annotation) that is not resolved in known structures of this protein. It seems to be located in the intracellular region (see Figure 9C), possibly disrupting interaction with proteins such as the calmodulin. Interestingly, PATHOS is predicting this mutation as pathogenic with a predicted score of 0.88, but not AlphaMissense which has a predicted score of 0.43 (see Figure 9B). This result demonstrates the interest and the efficiency of PATHOS to detect more disease-causing mutations than AlphaMissense, thanks to the richness of integrated features.

## Discussion

This study demonstrates that actual PLMs are worth using for the pathogenicity prediction task and a small combination of them is more beneficial than using all of them, saving a significant amount of computational time. We combined two PLMs, namely ESMC 600M and Ankh2 Large to build our VEP, called PATHOS, and show that it is among state of the art VEPs achieving excellent performance on most metrics and datasets.

Our results show that fine-tuning of these PLMs didn’t necessarily improve their performance on the validation dataset. Despite the fact that Rost Lab showed that fine-tuning PLM could increase performance (Schmirler et al., 2024), it does not seem to be the case for pathogenicity tasks, at least with our datasets. It could be interesting to see other fine-tuning of these PLMs with different datasets and verify if it comes from a data issue rather than an approach one. The decrease in performance may be the consequence of knowledge forgetting during the fine-tuning, despite the applied LoRA approach, which should avoid such events. More analysis needs to be done on the performance of fine-tuned PLMs, but our results were not sufficiently good to keep these refined models from original ones.

We consider that PATHOS cannot be classified as a meta-method like MetaRNN or ClinPred even if multiple algorithms were combined. Indeed, components of PATHOS were all trained on the same dataset (and are actually not VEPs), in contrast to MetaRNN for example, which combines 16 existing VEPs in its algorithm. The main difference lies in the ability to have full control on possible biases of each component of the VEP, which can not be done for classical meta-methods. They combine already trained VEPs without the possibility to adjust their training dataset. Both combinations have their strengths and weaknesses, but a controlled combination, such as PATHOS and based on the results presented above, is a more robust and reliable approach to build a unified tool. In this specific case, the two PLMs composing PATHOS were trained on a meticulously processed dataset. This involved removing 76% of the initial data to eliminate type 1 and type 2 data circularity bias. This bias control allows for building a model that captures fundamental characteristics of pathogenic and benign variants, rather than biases inherent to clinical datasets.

PATHOS was evaluated on three clinical datasets that come from our previous benchmark analysis (Radjasandirane et al., 2025). These datasets were processed to remove as much as possible biases such as data circularity type 1 and type 2, in order to make a fair evaluation of VEPs. Using robust metrics such as MCC, F1 score and the rank score, our results demonstrate that PATHOS is one of the best VEPs among the 66 evaluated VEPs. An improved MCC and F1 score is key in clinical application as they inform about the ability of the model. Indeed, it correctly classifies true positive and true negative while avoiding over predicting one class and making predictions errors. The Area Under the Receiver Operating Characteristic curve (AUROC) metric has not been considered in this study, as it often overestimates model performance. As explained in this paper (Lobo et al., 2008), AUROC mainly assesses the performance of the model in correctly classifying positive labels but does not include other important aspects of a model performance, such as precision or other metrics extracted from the confusion matrix, whereas the MCC does. A high MCC value corresponds unavoidably to a high confusion matrix values, leading to high values for metrics based on this matrix such as sensitivity, specificity, precision and negative predictive value,. This makes the MCC the gold standard metric to evaluate a model performance on a binary classification task. Based on these assumptions and on observed MCC performances, PATHOS can be considered among state of the art VEPs for clinical pathogenicity prediction. Indeed, it achieved the highest MCC on almost all datasets, including on ‘new’ sequences, indicating a higher and more robust quality of performance compared to evaluated VEPs.

Similarly to the MCC metric, the F1 score also combines multiple metrics as it is defined as the harmonic mean of the precision and recall metrics, therefore making the F1 score an highly relevant metric to estimate performance of a model in binary classification task. Especially, an high F1 score indicates that the model correctly predicts true positive while avoiding false positive and false negative prediction. In a clinical context, this means that the model can correctly find pathogenic variants while not overpredicting pathogenicity on benign variants, which is key for a patient harboring many mutations with only a few or one responsible for the observed disease. PATHOS has the highest F1 score (or a statistically non-significant rank from first VEP) on all datasets making PATHOS as one of the most reliable VEP for detecting pathogenic variants without overprediction. The rank score metric is the third main metric used to evaluate PATHOS performance directly against other 65 VEPs using shared variants. PATHOS achieved the highest rank score for all datasets, indicating that it significantly performs better or similarly to 87.7%, 95.4% and 80% of VEPs on respectively the Clinical, ClinVar and ClinVar_HQ datasets (see Figure 4C, 5C and 5F). This performance remains high when using only new sequences, demonstrating that PATHOS has excellent generalization (see Figure 6 and S8). Computing the mean rank score of ClinVar_HQ and Clinical datasets (high quality clinical variants) results in a relatively high averaged score of 0.838, outperforming AlphaMissense on the overall performance (see Figure S6).

The BER values for all datasets are relatively low (see Figure S9 A, B and C). PATHOS is consistently among the top 5 for BER, indicating a good balance between sensitivity and specificity metrics for each dataset. Indeed, PATHOS is not necessarily always among the top VEPs concerning sensitivity (see Figure S10 A, B and C) and specificity (see Figure S10 D, E and F), but the VEPs that rank ahead of PATHOS have an imbalance between sensitivity and specificity. For example, MetaRNN, which appears to have a sensitivity of 0.921 on Clinical (see Figure S10A) but has a specificity of 0.42 (see Figure S10D).

PATHOS is competitive in correctly predicting hard variants annotations, beating AlphaMissense on all datasets. The distinction between easy and hard variants can be done using the solvent accessibility on the protein structure as done in (Radjasandirane et al., 2025). However, no improvement has been observed when adding this feature into the model (data not shown), which may suggest that embedding information from PLMs is already incorporating such structural information.

Our analysis demonstrated that PATHOS is able to better detect functionally important regions within the sequence compared to AlphaMissense. Results presented in Figure 8 show that PATHOS did not overpredict pathogenicity on the region of the third known motif for PGR near residue 185. Instead, PATHOS seems to correctly recognize the sequence context to evaluate its biological importance and relevance. This result is consistent with the absence of clear evidence of any functional role of this motif in UniProt, in contrast to the first two LXXL motifs for which PATHOS correctly identifies the sequence as biologically relevant. Furthermore, analyzing the potassium channel KCNQ1 showed that PATHOS can find disease-causing variants that AlphaMissense misses like the mutation T587M. Such pathogenicity heatmaps can be helpful for clinical professionals. Many patients harbor Variants of Uncertain Significance (VUS) that have no clear annotations in public clinical databases and represent a key challenge. The clinician can eventually localize the VUS position on the heatmap and if it falls on a pathogenicity patch detected by PATHOS (or another VEP), it can be a preliminary clue regarding its potential pathogenic impact, thus contributing to the diagnostic pathway.

Finally, we show that PATHOS is a versatile tool that deserves to be among state-of-the-art VEPs for clinical pathogenicity interpretation, as it performs very well on multiple different metrics. A webserver has been developed to facilitate its usage for non-specialists. We are also releasing precomputed predictions for 140 million mutations from human proteins in the Swiss-Prot sub-database, facilitating access to prediction scores without needing to perform additional calculations. Another major contribution of this work is making the calculated embeddings (for both PLMs) available for 140 millions missense mutations in the human proteome. This value will continuously grow to attain all possible mutations on Swiss-Prot (around 200 millions). Generating such a quantity of embeddings requires significant resources, notably the use of several H100s in parallel (representing about 4000 GPU hours). By providing access to this data, we hope to remove a significant technical barrier limiting research groups with more modest means. We are confident that this data will help accelerate mutation-related research, which may lead to the development of more effective methods. Thus, any new method developed from these embeddings will be able to provide pre-computed predictions for the human proteome much more easily. The most time and resource intensive step having already been accomplished, this radically simplifies the use of future PLM-based VEPs, making them more accessible to non-specialists.

Although PATHOS performs very well, there are some factors that could limit its performance. First, its dependence on features like allele frequency and STRING interaction scores introduces challenges with missing data. Default values are assigned when a variant has missing data for these features, but this imputation strategy could potentially decrease prediction quality for concerned variants. Second, PATHOS uses features derived from UniProt annotations, which are based on the canonical protein sequence. This means PATHOS is not applicable for analyzing variants found on protein isoforms or other non-canonical sequences. Finally, the integration of two PLMs can make PATHOS computationally demanding for *de novo* predictions. We tried to address this issue by precomputing predictions for 140 million human missense mutations on Swiss-Prot but this aspect of PATHOS can eventually limit its applicability on a very large de novo dataset.

## Data Availability

Evaluation datasets used in this study can be accessed at https://github.com/DSIMB/VEP-Benchmark.

## Acknowledgments

The authors would like to thank Mr. Yann Vander Meersche and Pr. Catherine Etchebest (DSIMB, Paris, France) for fruitful discussions, as well as IDRIS for providing computational resources, including access to A100 GPUs (Allocation number: AD011014139) and H100 GPUs (Allocation number: AD010316397, used to infer embeddings on 140 millions mutations).

## Author contributions

A.G.dB. and J-C.G have designed and conceptualized the study. R.R. has made the curation of data and all the calculations. Analysis has been made by R.R. all the results have been discussed and improvements have been proposed by all authors. G.C. participated with conceptual insights. R.R. draws the initial draft and A.G. dB., J-C.G. and J.D. have edited the article. All authors have acknowledged this final version of the manuscript.

## Data and code availability

The stand-alone program is available at https://github.com/DSIMB/PATHOS, and the web server is freely accessible at https://dsimb.inserm.fr/PATHOS. Evaluation datasets used in this study can be accessed at https://github.com/DSIMB/VEP-Benchmark.

