## Supplementary materials for "PATHOS: Predicting Variant Pathogenicity by Combining Protein Language Models and Biological Features"

**Supplementary figures**

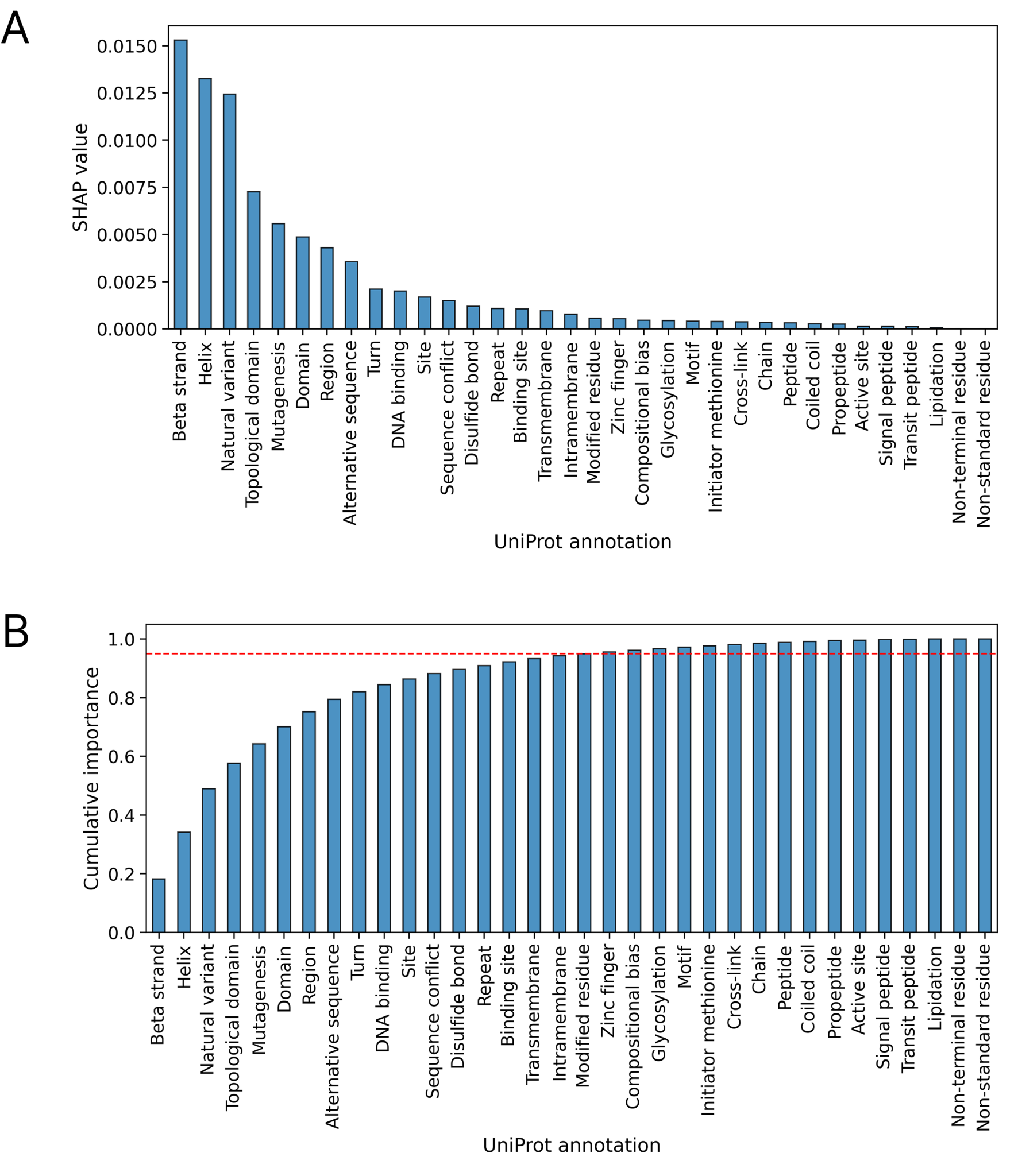

**Figure S1: SHAP value and cumulative sum computed for each UniProt annotation.** (A) Histogram of SHAP value. (B) The cumulative sum of SHAP values. The red dashed line represents the 95% cumulative importance threshold used to retain the most influential features.

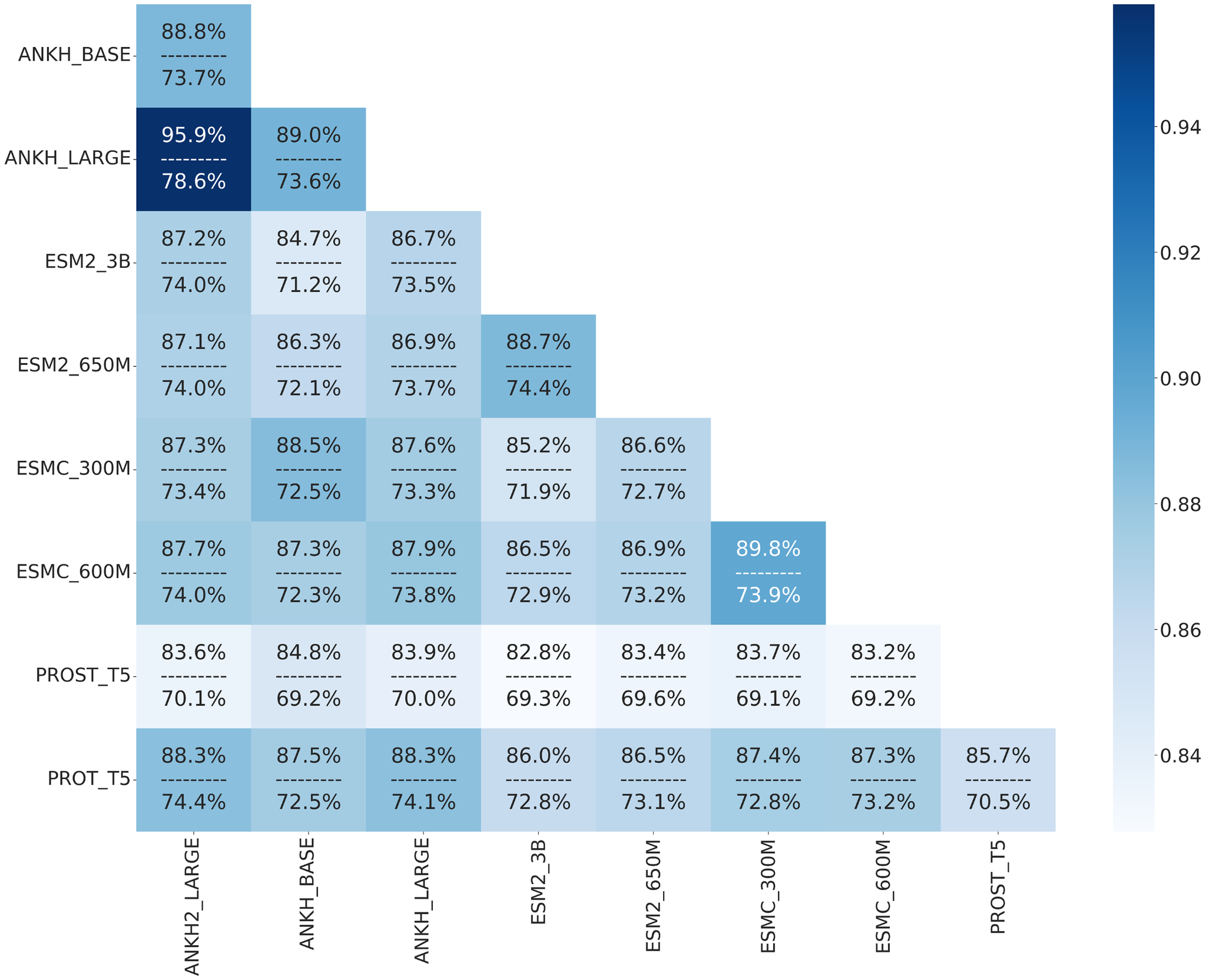

**Figure S2: Redundancy analysis of PLMs predictions.** This heatmap visualizes the redundancy and predictive agreement between different PLMs. For each pair of PLMs, the top value represents the percentage of identical predictions made by both models. The bottom value indicates the percentage of correct predictions among these identical predictions. The color intensity reflects the proportion of identical predictions, as shown by the color bar.

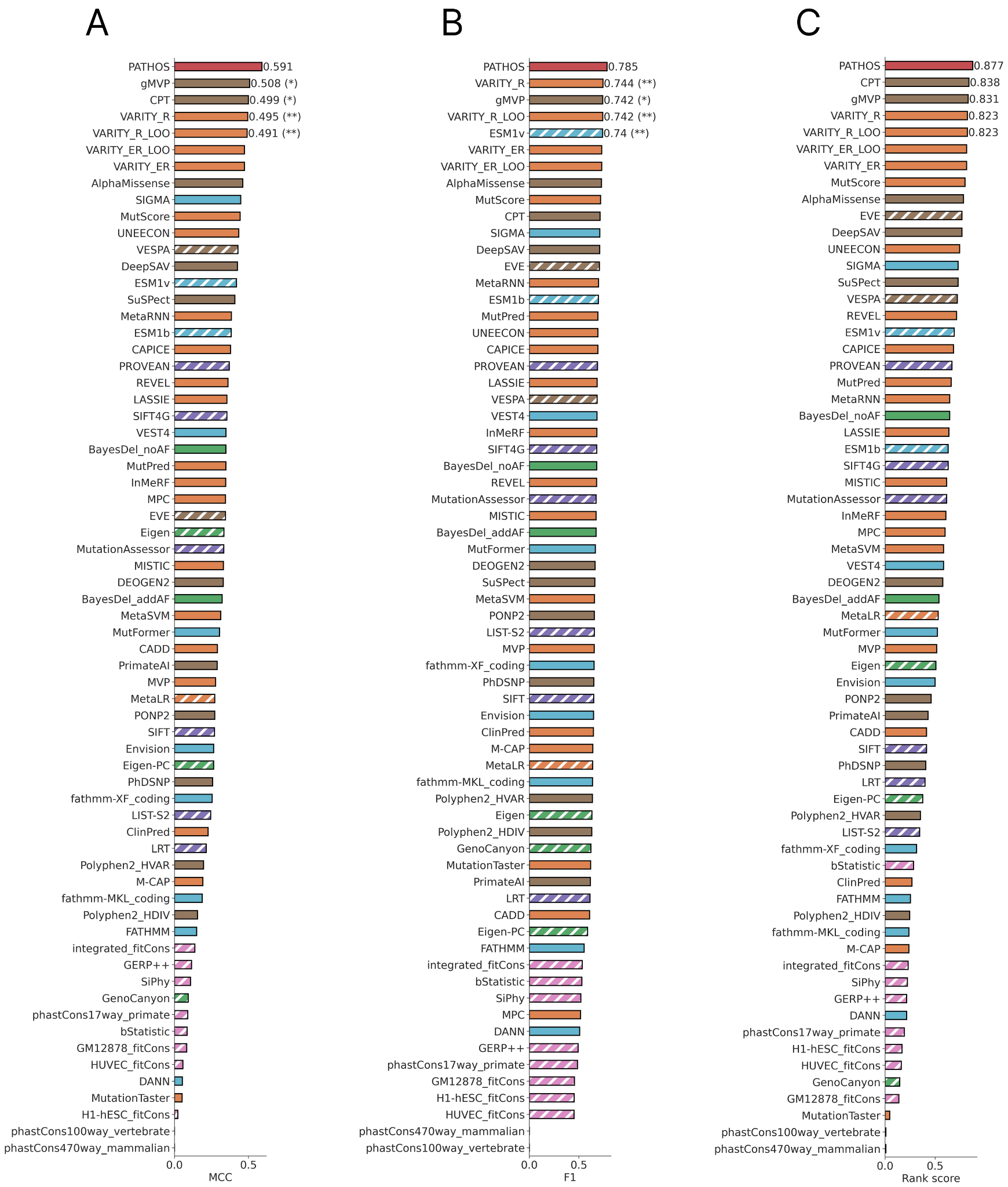

**Figure S3: Performance of 66 VEPs on Clinical dataset.** Histogram of MCC (A), F1 (B) and Rank score (C) values for the Clinical datasets. Each VEP is colored based on its algorithm type used as depicted by the legend on the bottom of the figure. Statistical differences between PATHOS and top-ranked VEPs have been assessed using the bootstrap method on 10,000 iterations.

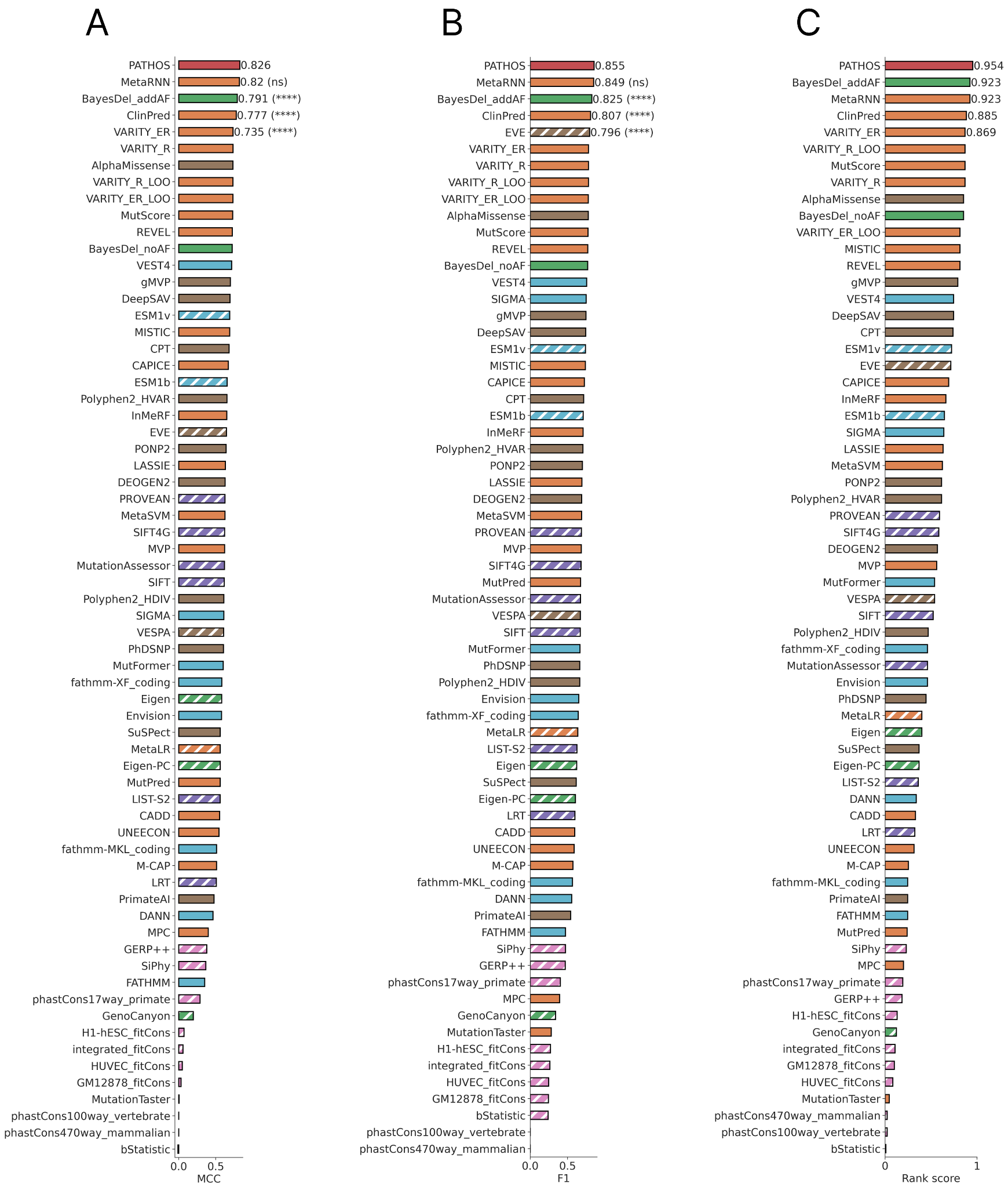

**Figure S4: Performance of 66 VEPs on ClinVar dataset.** Histogram of MCC (A), F1 (B) and Rank score (C) values for the ClinVar dataset. Each VEP is colored based on its algorithm type used as depicted by the legend on the bottom of the figure. Statistical differences between PATHOS and top-ranked VEPs have been assessed using the bootstrap method on 10,000 iterations.

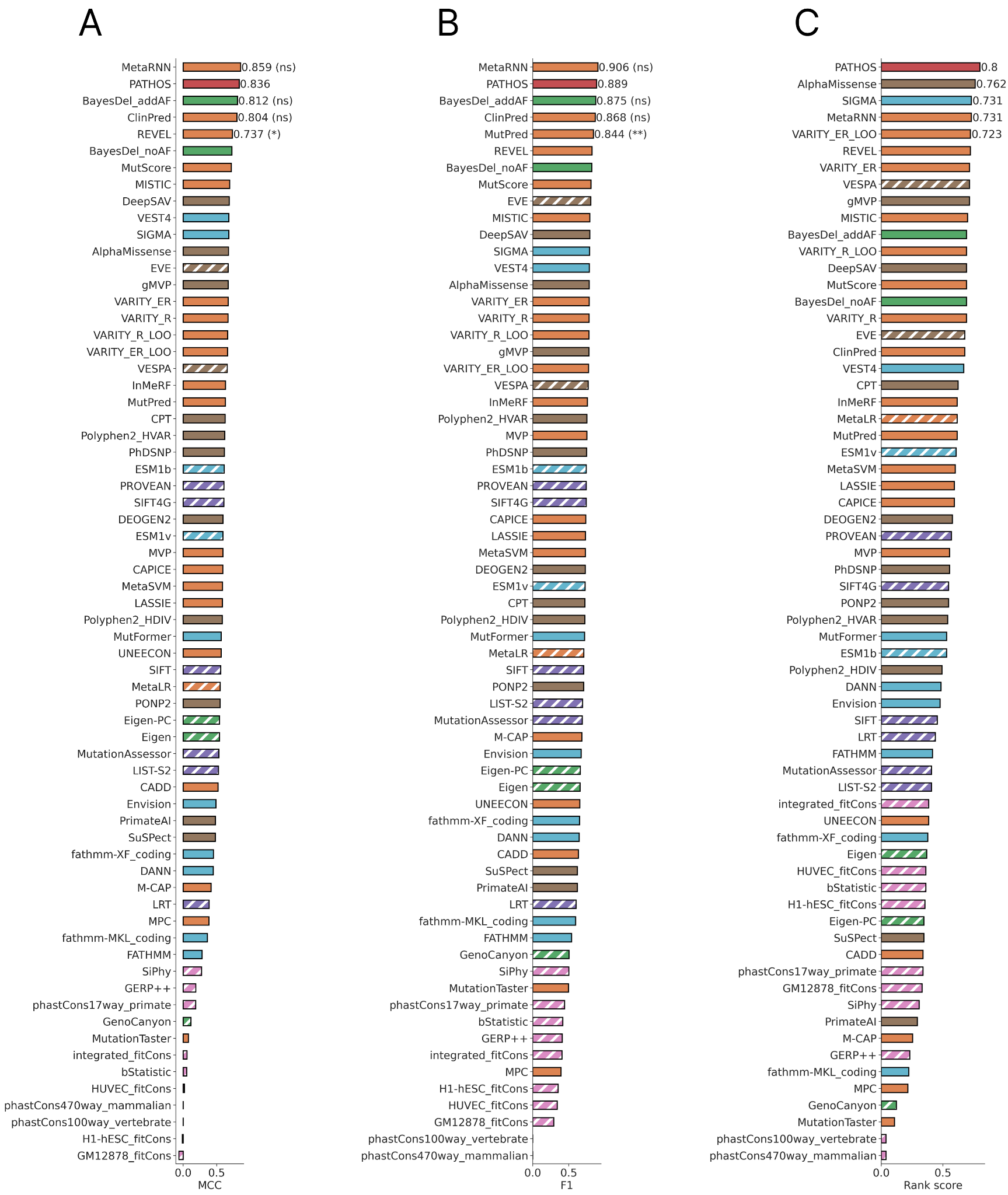

**Figure S5: Performance of 66 VEPs on ClinVar_HQ dataset.** Histogram of MCC (A), F1 (B) and Rank score (C) values for the ClinVar_HQ dataset. Each VEP is colored based on its algorithm type used as depicted by the legend on the bottom of the figure. Statistical differences between PATHOS and top-ranked VEPs have been assessed using the bootstrap method on 10,000 iterations.

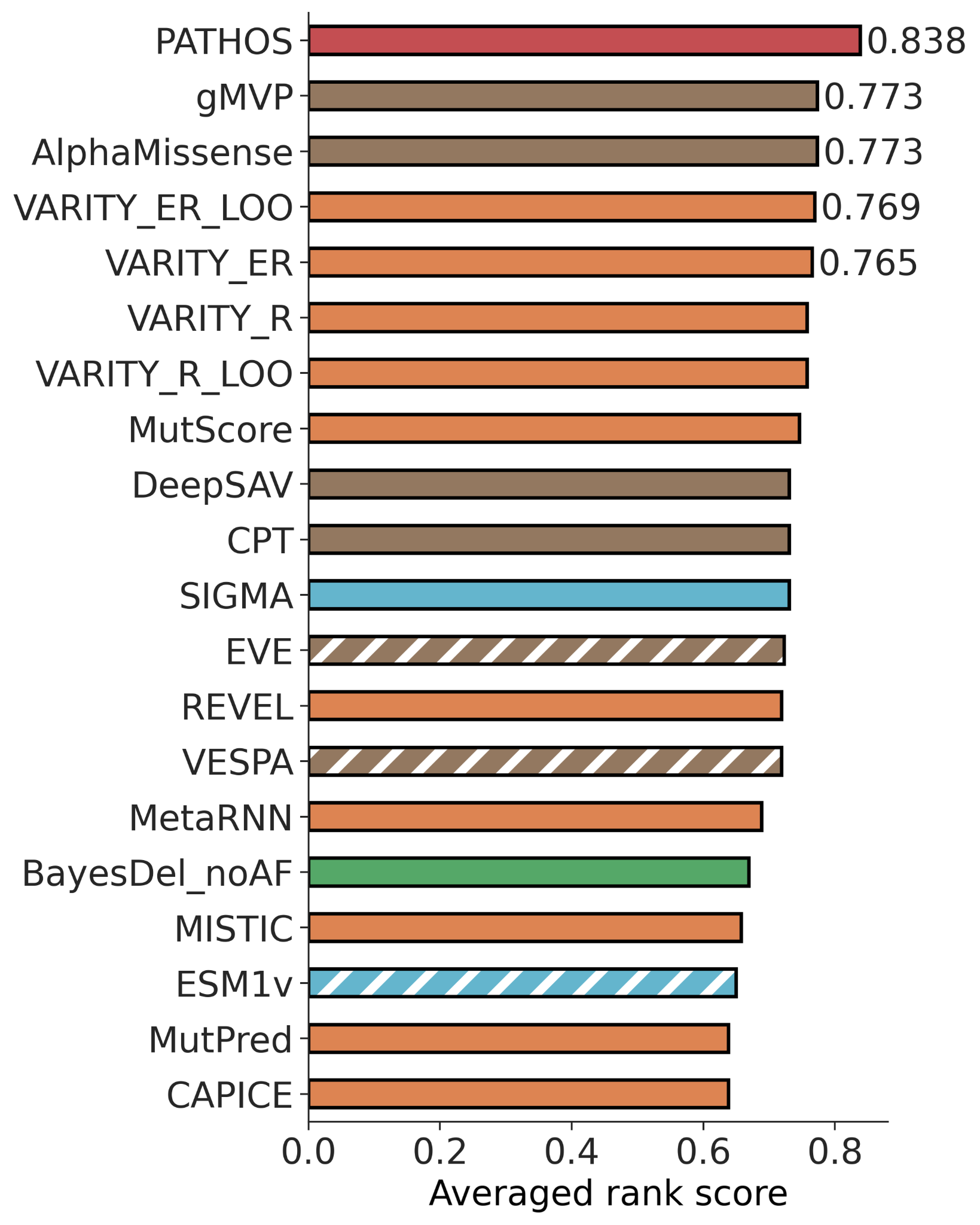

**Figure S6: Averaged performance on clinical variants.** The average rank score represents the mean between rank scores values from ClinVar_HQ and Clinical datasets and constitute the global performance of each VEP on clinical variants

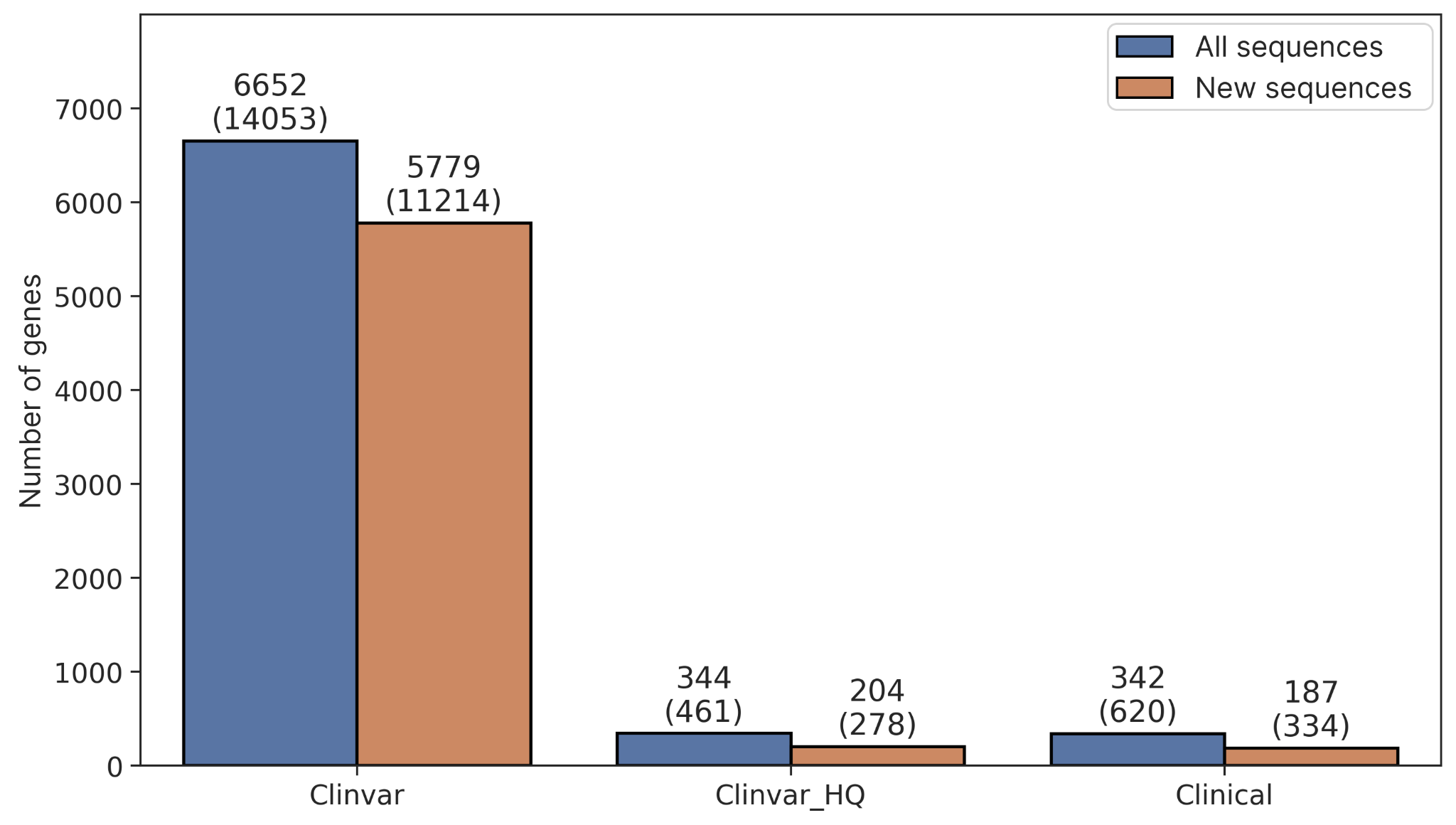

**Figure S7: Number of genes and variants among new sequences in each dataset.** Each bar represents the number of genes, with the gene count value above the bars and the corresponding variant count in parentheses. The blue bars relate to the initial datasets, and the orange bars relate to the datasets containing only the new sequences.

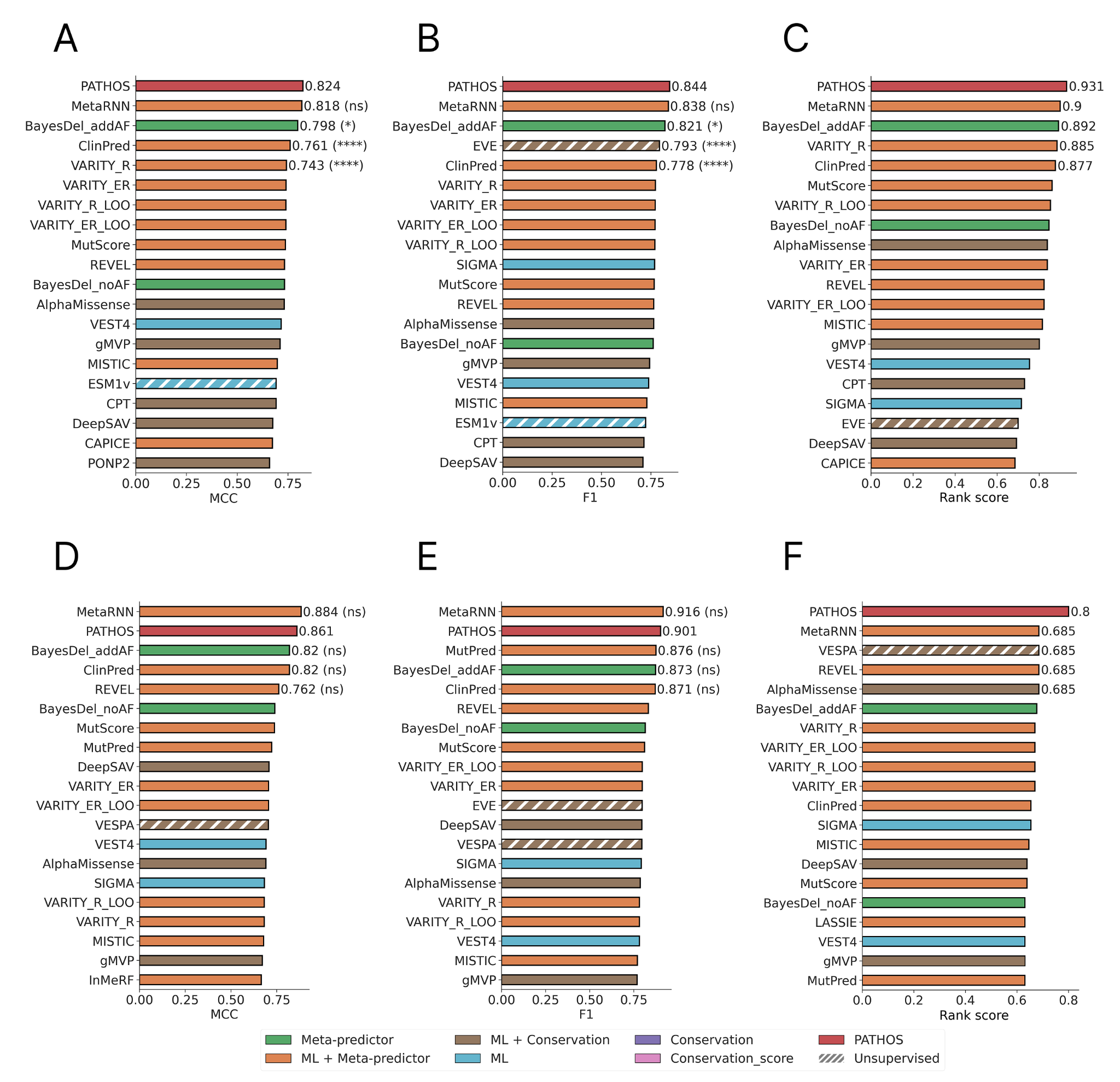

**Figure S8: Performance on ClinVar and ClinVar_HQ dataset using new sequences absent from the training dataset.** Histogram of MCC (A), F1 (B) and Rank score (C) values for the ClinVar datasets, and MCC (D), F1 (E) and Rank score (F) for the ClinVar_HQ dataset. Only the top 20 VEPs out of 66 are shown. Each VEP is colored based on its algorithm type used as depicted by the legend on the bottom of the figure. Statistical differences between PATHOS and top-ranked VEPs have been assessed using the bootstrap method on 10,000 iterations.

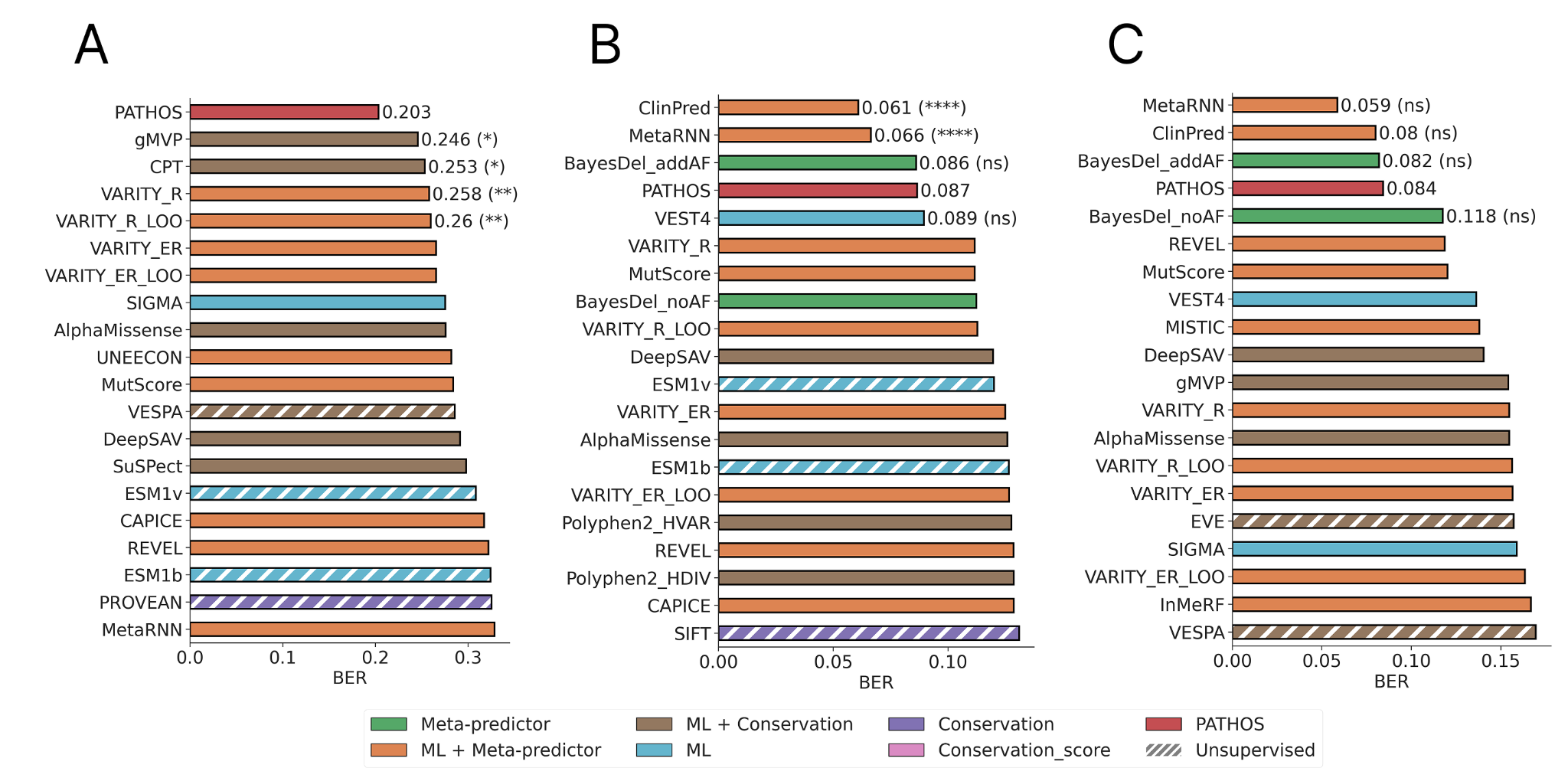

**Figure S9: BER value on each dataset.** Bar plot of BER values for (A) Clinical, (B) ClinVar, and (C) ClinVar_HQ. Only the top 20 VEPs out of 66 are shown. Each VEP is colored according to its algorithm type, as indicated in the legend at the bottom of the figure. Statistical differences between PATHOS and the top-ranked VEPs were assessed using the bootstrap method over 10,000 iterations.

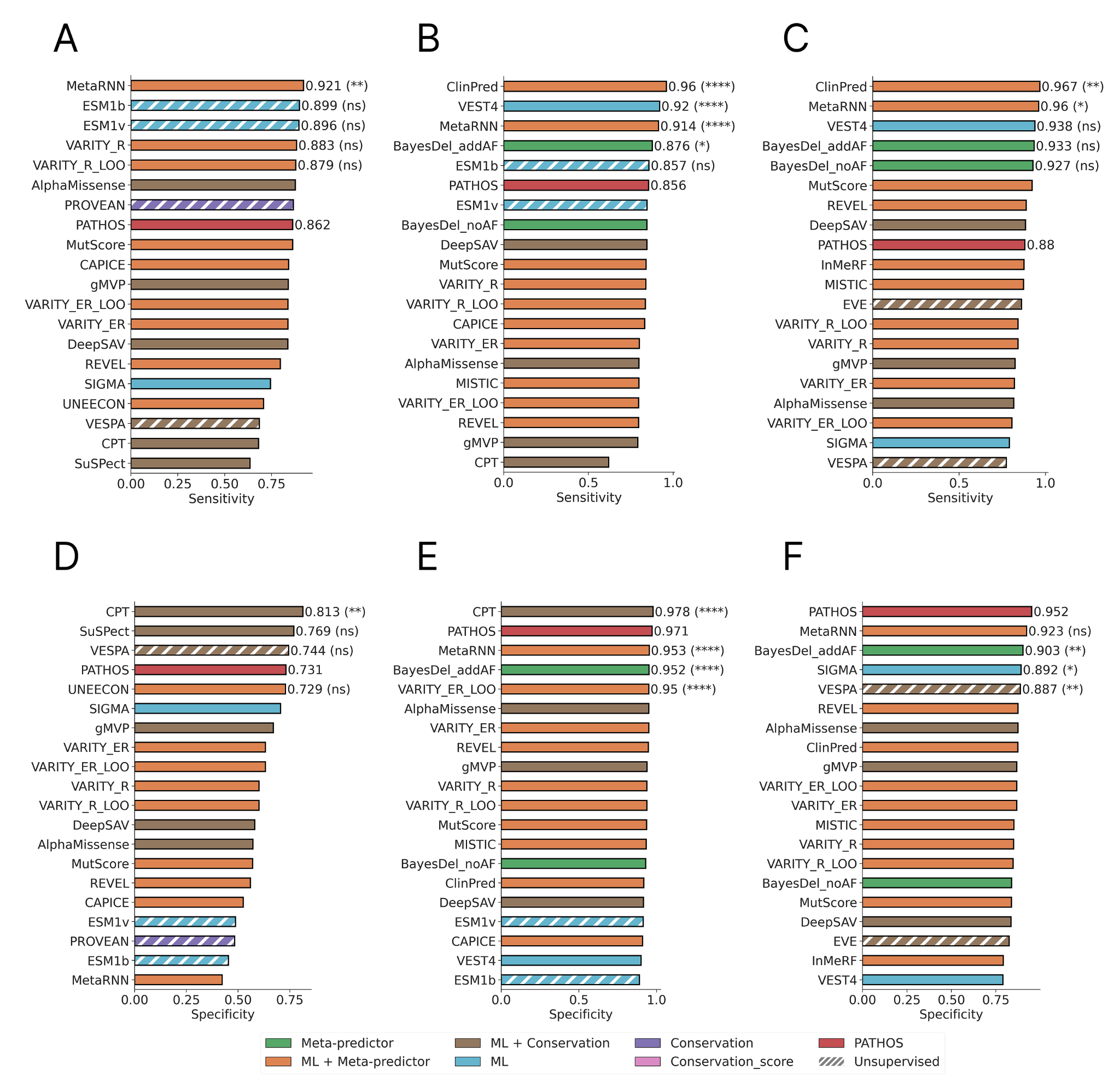

**Figure S10: Sensitivity and specificity values on each dataset.** Bar plot of sensitivity values for (A) Clinical, (B) ClinVar, and (C) ClinVar_HQ, and specificity values for (D) Clinical, (E) ClinVar, and (F) ClinVar_HQ. Only the top 20 VEPs out of 66 according to MCC are presented, in order to avoid showing VEPs with a sensitivity of 1 but low specificity. Each VEP is colored according to its algorithm type, as indicated in the legend at the bottom of the figure. Statistical differences between PATHOS and the top-ranked VEPs were assessed using the bootstrap method over 10,000 iterations.

**Supplementary tables**

| Model | Hugging Face repo name | Revision version |
| --- | --- | --- |
| Ankh2 Large | ElnaggarLab/ankh2-ext2 | 4c155ee6b1aeb7f29ebf06a0399b331504104b67 |
| Ankh Large | ElnaggarLab/ankh-large | 74b371dbfa3ee0a05d32ae74df0c2e0b82d6b9a6 |
| Ankh Base | ElnaggarLab/ankh-base | d99cb6b966530dfc2ae96bc69d9255c2a07308b0 |
| ESM2 3B | facebook/esm2_t36_3B_UR50D | 476b639933c8baad5ad09a60ac1a87f987b656fc |
| ESM2 650M | facebook/esm2_t33_650M_UR50D | 08e4846e537177426273712802403f7ba8261b6c |
| ProtT5 | Rostlab/prot_t5_xl_uniref50 | 973be27c52ee6474de9c945952a8008aeb2a1a73 |
| ProstT5 | Rostlab/ProstT5 | d7d097d5bf9a993ab8f68488b4681d6ca70db9e5 |
| ESMC 600M | Synthyra/ESMplusplus_large | 1408244c8c08fb1b593da75b912b56a1688be86e |
| ESMC 300M | Synthyra/ESMplusplus_small | e655f5a1ccd6062cd99a0435ba83672639e18593 |

**Table S1: Hugging Face repo name and version used for each PLM.** The table shows PLM versions used for this study. Using a different version from these ones can lead to different results as they have not been used for our training.

| Features | Description |
| --- | --- |
| PLMs embeddings | Embedding vector of the mutant and the wildtype amino acid.  PATHOS incorporates two PLMs namely ESMC 600M and Ankh2 Large. |
| PastML | Probability to observe the mutation based on the evolution history of the mutated position. |
| Allele Frequency | Frequency extracted from gnomAD database if available. The default value is 0. |
| STIRING | Propensity of a protein to interact, calculated from the STRING database.  The default value is the average of the normalized STRING score of the training set, which is 0.583781963271417. |
| UniProt annotations | The UniProt annotation feature is represented as a 34-dimensional binary vector. For a given protein ID, a '1' indicates the presence and a '0' indicates the absence of a specific annotation type within a window of plus or minus 5 residues around the mutated position.  The possible annotation from the UniProt GFF file are:  -Active site (X)  **-Alternative sequence**  **-Beta strand**  **-Binding site**  -Chain (X)  -Coiled coil (X)  -Compositional bias (X)  -Cross-link (X)  **-DNA binding**  **-Disulfide bond**  **-Domain**  -Glycosylation (X)  **-Helix**  -Initiator methionine (X)  **-Intramembrane**  -Lipidation (X)  **-Modified residue**  -Motif (X)  **-Mutagenesis**  **-Natural variant**  -Non-standard residue (X)  -Non-terminal residue (X)  -Peptide (X)  -Propeptide (X)  **-Region**  **-Repeat**  **-Sequence conflict**  -Signal peptide (X)  **-Site**  **-Topological domain**  -Transit peptide (X)  **-Transmembrane**  **-Turn**  -Zinc finger (X) |

**Table S2: Features used in the PATHOS model.** The table shows features described in the section 5 of Material and Methods section. UniProt annotations with a ‘X’ are not incorporated in the model, those in bold are used to train PATHOS.

| **VEP** | **Year** | **Type** | **Approach** | **Nb of scores** | **Threshold** | **Input type** | **Source** | **Reference** |
| --- | --- | --- | --- | --- | --- | --- | --- | --- |
| SIFT | 2003 | Conservation | Unsupervised | 1 | 0.05 | Genomic position | dbNSFP | (Ng & Henikoff, 2003) |
| phastCons | 2005 | Conservation score | Unsupervised | 3 | 1 | Genomic position | dbNSFP | (Siepel et al., 2005) |
| Siphy | 2009 | Conservation score | Unsupervised | 1 | 15.47 | Genomic position | dbNSFP | (Garber et al., 2009) |
| bStatistic | 2009 | Conservation score | Unsupervised | 1 | 797.5 | Genomic position | dbNSFP | (McVicker et al., 2009) |
| LRT | 2009 | Conservation | Unsupervised | 1 | 0.000005 | Genomic position | dbNSFP | (Chun & Fay, 2009) |
| MutPred | 2009 | ML + Meta-predictor | Supervised | 1 | 0.5 | Genomic position | dbNSFP | (B. Li et al., 2009) |
| GERP++ | 2010 | Conservation score | Unsupervised | 1 | 5.26 | Genomic position | dbNSFP | (Davydov et al., 2010) |
| Polyphen2 | 2010 | ML + Conservation | Supervised | 2 | 0.5 | Genomic position | dbNSFP | (Adzhubei et al., 2010) |
| MutationTaster | 2010 | ML + Meta-predictor | Supervised | 1 | 0.5 | Genomic position | dbNSFP | (Schwarz et al., 2010) |
| MutationAssessor | 2011 | Conservation | Unsupervised | 1 | 1.935 | Genomic position | dbNSFP | (Reva et al., 2011) |
| PROVEAN | 2012 | Conservation | Unsupervised | 1 | -2.5 | Genomic position | dbNSFP | (Choi et al., 2012) |
| VEST4 | 2013 | ML | Supervised | 1 | 0.5 | Genomic position | dbNSFP | (Carter et al., 2013) |
| FATHMM | 2013 | ML | Supervised | 1 | -1.5 | Genomic position | dbNSFP | (Shihab et al., 2013) |
| SuSPeCT | 2014 | ML + Conservation | Supervised | 1 | 50 | Protein position | Local execution | (Yates et al., 2014) |
| DANN | 2014 | ML | Supervised | 1 | 1 | Genomic position | dbNSFP | (Quang et al., 2015) |
| CADD | 2014 | ML + Meta-predictor | Supervised | 1 | 3.957 | Genomic position | dbNSFP | (Kircher et al., 2014) |
| MetaLR | 2015 | ML + Meta-predictor | Unsupervised | 1 | 0.5 | Genomic position | dbNSFP | (Dong et al., 2015) |
| MetaSVM | 2015 | ML + Meta-predictor | Supervised | 1 | 0 | Genomic position | dbNSFP | (Dong et al., 2015) |
| fitCons | 2015 | Conservation score | Unsupervised | 4 | 0.676 | Genomic position | dbNSFP | (Gulko et al., 2015) |
| PONP2 | 2015 | ML + Conservation | Supervised | 1 | 0.5 | Protein position | Precomputed | (Niroula et al., 2015) |
| FATHMM-MKL | 2015 | ML | Supervised | 1 | 0,5 | Genomic position | dbNSFP | (Shihab et al., 2015) |
| GenoCanyon | 2015 | Meta-predictor | Unsupervised | 1 | 0.5 | Genomic position | dbNSFP | (Lu et al., 2015) |
| SIFT4G | 2016 | Conservation | Unsupervised | 1 | 0.05 | Genomic position | dbNSFP | (Vaser et al., 2016) |
| BayesDel | 2016 | Meta-predictor | Supervised | 2 | 0.0692; -0.0570 * | Genomic position | dbNSFP | (Feng, 2017) |
| Eigen | 2016 | Meta-predictor | Unsupervised | 2 | 0.685 | Genomic position | dbNSFP | (Ionita-Laza et al., 2016) |
| M-CAP | 2016 | ML + Meta-predictor | Supervised | 1 | 0.025 | Genomic position | dbNSFP | (Jagadeesh et al., 2016) |
| REVEL | 2016 | ML + Meta-predictor | Supervised | 1 | 0.5 | Genomic position | dbNSFP | (Ioannidis et al., 2016) |
| DEOGEN2 | 2017 | ML + Conservation | Supervised | 1 | 0.5 | Genomic position | dbNSFP | (Raimondi et al., 2017) |
| PhDSNP | 2017 | ML + Conservation | Supervised | 1 | 0.5 | Genomic position | Local execution | (Capriotti & Fariselli, 2017) |
| MPC | 2017 | ML + Meta-predictor | Supervised | 1 | 2 | Genomic position | dbNSFP | (Samocha et al., 2017) |
| PrimateAI | 2018 | ML + Conservation | Supervised | 1 | 0.803 | Genomic position | dbNSFP | (Sundaram et al., 2018) |
| FATHMM-XF | 2018 | ML | Supervised | 1 | 0,5 | Genomic position | dbNSFP | (Rogers et al., 2018) |
| ClinPred | 2018 | ML + Meta-predictor | Supervised | 1 | 0.5 | Genomic position | dbNSFP | (Alirezaie et al., 2018) |
| Envision | 2019 | ML | Supervised | 1 | 0.9 | Protein position | Precomputed | (Gray et al., 2018) |
| LASSIE | 2019 | ML + Meta-predictor | Supervised | 1 | 0.0005 | Genomic position | Precomputed | (Huang & Siepel, 2019) |
| LIST-S2 | 2020 | Conservation | Unsupervised | 1 | 0.85 | Genomic position | dbNSFP | (Malhis et al., 2020) |
| DeepSAV | 2020 | ML + Conservation | Supervised | 1 | 0.44 | Protein position | Precomputed | (Pei et al., 2020) |
| InMeRF | 2020 | ML + Meta-predictor | Supervised | 1 | 0.5 | Genomic position | Precomputed | (Takeda et al., 2020) |
| MISTIC | 2020 | ML + Meta-predictor | Supervised | 1 | 0.5 | Genomic position | Precomputed | (Chennen et al., 2020) |
| CAPICE | 2020 | ML + Meta-predictor | Supervised | 1 | 0.02 | Genomic position | Precomputed | (S. Li et al., 2020) |
| UNEECON | 2020 | ML + Meta-predictor | Supervised | 1 | 0.5 | Genomic position | Precomputed | (Huang, 2020) |
| EVE | 2021 | ML + Conservation | Unsupervised | 1 | 0.5 | Protein position | Precomputed | (Frazer et al., 2021) |
| ESM1v | 2021 | ML | Unsupervised | 1 | -7.5 | Protein position | Local execution | (Meier et al., 2021) |
| MutFormer | 2021 | ML | Supervised | 1 | 0.5 | Genomic position | Precomputed | (Jiang et al., 2023) |
| MVP | 2021 | ML + Meta-predictor | Supervised | 1 | 0.75 | Genomic position | dbNSFP | (Qi et al., 2021) |
| VARITY | 2021 | ML + Meta-predictor | Supervised | 4 | 0.5 | Protein position | Precomputed | (Wu et al., 2021) |
| gMVP | 2022 | ML + Conservation | Supervised | 1 | 0.75 | Genomic position | dbNSFP | (Zhang et al., 2022) |
| VESPA | 2022 | ML + Conservation | Unsupervised | 1 | 0.5 | Protein position | Precomputed | (Marquet et al., 2022) |
| MetaRNN | 2022 | ML + Meta-predictor | Supervised | 1 | 0.5 | Genomic position | dbNSFP | (C. Li et al., 2022) |
| MutScore | 2022 | ML + Meta-predictor | Supervised | 1 | 0.5 | Genomic position | Precomputed | (Quinodoz et al., 2022) |
| CPT | 2023 | ML + Conservation | Supervised | 1 | 0.5 | Protein position | Precomputed | (Jagota et al., 2023) |
| AlphaMissense | 2023 | ML + Conservation | Supervised | 1 | 0.5 | Protein position | Precomputed | (Cheng et al., 2023) |
| ESM1b | 2023 | ML | Unsupervised | 1 | -7.5 | Protein position | Precomputed | (Brandes et al., 2023) |
| SIGMA | 2024 | ML | Supervised | 1 | 0.5 | Genomic position | Precomputed | (Zhao et al., 2024) |

**Table S3. List of VEPs evaluated in this study.** For each VEP is provided its year of publication, the type of the algorithm (6 classes are defined: Conservation, ML, Conservation + ML, Meta-predictor, ML + Meta-predictor, Conservation score, see section Groups of VEP) and approach (supervised or not), the number of scores provided, the threshold used and the source where predictions were retrieved. These last can come from three possible sources. (i) dbNSFP, predictions are coming from the database of prediction dbNSFP, (ii) Precomputed predictions, prediction are coming from parsing files of precomputed predictions provided with the VEP, and (iii) Local run, prediction are coming from local run of the VEP.

* BayesDel has two versions and a different threshold for each: BayesDel_addAF 0.0692 and BayseDel_noAF -0.057.

|  |  |  | **Fine-tuning** | | | | **Non-fine-tuning** | | | |
| --- | --- | --- | --- | --- | --- | --- | --- | --- | --- | --- |
| Model | Embedding size | Number of Parameters | Number of trainable parameters (LoRA) | Time to pre-generate embeddings before training | Time to train the model | Epoch | Number of trainable parameters | Time to pre-generate embeddings before training (100,260 mutations) | Time to train the model | Epoch |
| Ankh2 Large | 1536 | 1,151,710,724 | 1,970,180 | - | 720 minutes | 1 | 1537 | 117.6 minutes | 10.0 minutes | 43 |
| Ankh Large | 1536 | 1,151,710,724 | 1,970,180 | - | 720 minutes | 1 | 1537 | 119.3 minutes | 16.0 minutes | 68 |
| Ankh Base | 768 | 453,172,232 | 1,181,960 | - | 450 minutes | 1 | 769 | 59.6 minutes | 6.8 minutes | 29 |
| ESM2 3B | 2560 | 3,000,000,000 | 7,656,965 | - | 420 minutes | 1 | 2561 | 263.8 minutes | 2.3 minutes | 10 |
| ESM2 650M | 1280 | 650,000,000 | 3,509,769 | - | 180 minutes | 1 | 1281 | 71.1 minutes | 2.8 minutes | 12 |
| ProtT5 | 1024 | 1,208,143,876 | 1,969,156 | - | 300 minutes | 1 | 1025 | 118.6 minutes | 5.3 minutes | 23 |
| ProstT5 | 1024 | 1,208,166,408 | 1,969,160 | - | 240 minutes | 1 | 1025 | 99.0 minutes | 4.6 minutes | 20 |
| ESMC 600M | 1152 | 600,000,000 | - | - | - | - | 1153 | 51.9 minutes | 6.5 minutes | 28 |
| ESMC 300M | 960 | 300,000,000 | - | - | - | - | 961 | 33.1 minutes | 9.6 minutes | 41 |

**Table S4: Comparison of PLMs parameters and training information between fine-tuning and non-fine-tuning approaches.** This table details key characteristics of the evaluated PLMs, including their embedding size, total number of parameters (including LoRA parameters), and training times
